# Proactive Community Case Management to improve care seeking for febrile illness in Sierra Leone: A community randomized controlled trial

**DOI:** 10.1101/2025.11.06.25339612

**Authors:** Joshua Yukich, Annie Arnzen, Maia Cullen, Alyssa J. Young, Annē Linn, Ruth Ashton, Thomas P. Eisele, Kim Lindblade, Lakoh Sulaiman, Bailah Molleh, Ginika Egesimba, Megan Littrell

## Abstract

**Background:** Prompt treatment seeking for febrile illness is a critical step in the care cascade for malaria. In wide areas of sub-Saharan Africa there are critical gaps in coverage of prompt care. Integrated Community Case Management (iCCM) has been shown to be a burden reducing intervention that can influence malaria transmission and reduce overall disease burden and increase care access. Studies have not determined whether the delivery of iCCM through passive or proactive means provide ancillary benefits such as sustainable improvements in care-seeking behavior. This study was designed to determine if ProCCM could deliver sustainable increases in care seeking behavior even after a short period of implementation had ended.

**Methods:** A three-arm community randomized trial was conducted in Pujehun district, Sierra Leone from May 2024 until it was interrupted by the U.S. government in January 2025. The trial compared communities with up to four rounds of ProCCM and stock out support, to stock out support and iCCM alone, or to only iCCM alone (Standard Practice). Forty-six Primary Health Care Units (PHCU) were grouped into 45 clusters and randomized (1:1:1) to one of the three arms using restricted randomization. Treatment seeking rates were measured using cross-sectional household surveys at baseline (before randomization) and at one month post intervention. A final survey was planned at four months after intervention end but was interrupted by U.S. Government actions.

**Findings:** In the first survey post intervention, 250 of 391 febrile children sought care promptly in the preceding two weeks. Household education, asset ownership, access to water, household construction and use of malaria prevention were similar across arms. There was a significantly higher rate of prompt care-seeking for febrile illness in groups exposed to ProCCM as compared to the other arms. Among the hardest to reach populations, odds of care seeking increased nearly 3x (O.R. 2.78 (95% C.I. 1.03-7.85) *p*=0.05). Post-intervention beliefs about the availability, quality and community norms of care-seeking for fever were all higher in the ProCCM arm relative to the controls.

**Interpretation:** ProCCM delivered in short periods can improve care seeking for febrile illness in hard-to-reach populations by modifying community perceptions of community health workers availability and competence, when drugs and diagnostics for malaria care are available. We were unable to assess the sustainability of these gains due to interruption of PMI funding.

**Funding:** USAID/PMI Funding through PATH/PMI Insights supported the original study and data collection. All analytic and post-data collection work supported by Tulane University.

## Background and Objectives

Malaria case management that includes testing and treatment of people experiencing signs and symptoms of illness is an essential strategy in the fight against malaria in many countries in Africa. The utilization and impact of these services is largely dependent on people seeking care when they or their children have fever. In many malaria-affected countries, trends in care-seeking have not seen the same marked improvements that have been observed in other malaria intervention coverage indicators (1). The barriers to care-seeking are well documented including the cost of health services, travel time and distance, and negative perceptions regarding the quality-of-care available (2–4). Despite knowledge of these barriers, no broad increase in care seeking for fever has resulted from general investments in health care strengthening in sub-Saharan Africa (1).

In recent years, many countries have introduced and scaled up community case management (CCM) of malaria (and, in many settings, integrated management of childhood illness or iCCM, which includes malaria, pneumonia, and diarrhea) through community health workers (CHWs). The availability of malaria services at the community level through CHWs addresses geographic barriers to care and increases access to life-saving services (5). However, the utilization of CHWs varies widely by context (1,6).

To reduce the gap in care-seeking and address some of the known barriers to seeking care and utilizing CHW services, proactive case finding strategies have been implemented in some countries to increase the prompt treatment of uncomplicated malaria and prevent such cases from developing into severe disease. One such strategy is proactive community case management (ProCCM). In this intervention, CHWs visit households in their communities to proactively offer a defined package of services, such as CCM for malaria or iCCM. In models of this intervention that focus on malaria, CHWs screen for persons with fever and offer malaria diagnostic and other assessment services, as well as malaria treatment, health communication, and referral services. Proactive community case management is implemented in addition to the CHWs’ routine activities providing passive CCM or iCCM in community settings. Studies of ProCCM have been conducted in a number of different countries and settings, with recent studies in Senegal (7,8), Madagascar (5), Mali (9,10), and Zambia (11).

ProCCM has been shown to be well accepted by the community, feasible to implement, and to increase the number of patients treated by CHWs (7,8). In some settings, the implementation of ProCCM has been linked to improvements in early care-seeking for fever (8). However, a systematic review conducted by Whidden et al found that while many studies of proactive case-finding home visits by CHWs increased access to care and reduced child mortality, there was no significant impact on care-seeking behavior (12). Additionally, studies of the introduction of CCM have shown that it can effectively reduce malaria burden but have failed to find significant differences in burden reduction based on the strategy used for CCM (*e.g.* proactive vs. passive).

Sierra Leone, where malaria remains the leading cause of illness and death among children under five years of age, first introduced a CHW program in 2012, which was then scaled nationwide in 2017. In addition to health prevention and promotional services, HTR CHWs are trained and supplied to provide expanded iCCM services, including diagnosis and treatment of malaria, diarrhea, and pneumonia. The Sierra Leone 2021 Malaria Indicator Survey found that advice or treatment was sought for 75 percent of children who had fever within the preceding two weeks, however only 40 percent sought care on the same or next day following fever onset (13). Despite the availability of CHWs, only 7 percent of caregivers who sought care for a child with recent fever reported seeking care from a CHW, with the majority seeking care from a government health facility (68 percent) (13).

Despite the critical importance of timely identification and treatment of malaria, many families do not promptly seek care for febrile illness for a variety of reasons. This study sought to determine whether, among communities living in HTR areas of Sierra Leone, a short period of ProCCM (specifically, four household visits in two months) conducted at the start of a peak malaria transmission season can improve early care-seeking for fever and whether these improvements are sustained for the duration of the malaria transmission season.

## Methods

### Trial Design

This study is a three-arm, cluster-randomized controlled trial conducted in Pujeun district in Sierra Leone. Clusters were constituted as the Hard-to-reach (HTR) communities in catchment areas of peripheral health units (PHUs) where at least two communities were classified as HTR and where there was at least one CHW trained and active in iCCM. The study arms received the following interventions:

Arm 1. ProCCM - Optimized standard of care for CCM plus ProCCM - ProCCM will be implemented for two months near the start of the high transmission season, and the existing program will be supplemented to mitigate stock outs of malaria commodities among HTR CHWs and ensure HTR CHWs are adequately trained on SBCC messages to ensure activities are implemented as designed.

Arm 2 – Stock Out Mitigation Only (*e.g.* Optimized standard of care – as in arm 1 without ProCCM) or iCCM plus Stock out Mitigation

Arm 3 - Routine implementation (control) – no changes (business as usual) to iCCM, SBCC and stock management.

Changes in care seeking were assessed through cross-sectional surveys. A baseline survey was conducted prior to randomization and intervention implementation followed by a second survey at or near the end of the ProCCM intervention period, and a final survey was planned at the end of the malaria transmission season.

### Participants

The target population for the intervention consisted of all *de facto* and *de jure* residents present in HTR communities in the intervention and control clusters during the study period and the CHWs who served these communities. The population to be sampled for outcome assessment included additional inclusion and exclusion criteria as outlined below.

### Interventions

In all three study arms, HTR CHWs will continue to provide the standard package of iCCM (passive case detection) plus services as described in the national CHW strategy. HTR CHWs typically serve a catchment population of 300 to 350 people (50 to 60 households) that is more than five kilometers from the nearest health facility or three to five kilometers in areas with particularly difficult terrain. HTR CHWs will be supervised by peer supervisors who have been trained and are in operation according to the national strategy. HTR CHWs typically serve only one community, in which they also reside, but occasionally they also serve adjacent communities meaning that not every HTR community will have a dedicated CHW though most did at the time of the trial.

### ProCCM

The ProCCM intervention was implemented at the beginning of the peak malaria transmission season for approximately two months. HTR CHWs and supervisors in communities assigned to this arm received additional training, supported by the national CHW hub, on the proactive intervention. CHWs were expected to visit each household in their catchment area twice per month for a total of four home visits per household. An additional monetary incentive was provided to HTR CHWs in this arm to compensate them for the extra effort and costs associated with adding the proactive visits to their existing scope of work.

During the visits, CHWs identified household members of all ages complaining of fever or history of fever and record household profile in a paper-based register. People with fever, or a history of fever in the past 48 hours, or other signs or symptoms suggestive of malaria were offered a rapid diagnostic test (RDT) for malaria. CHWs provided individuals that tested malaria positive with a course of antimalarials based on national guidance for dosing based on age and/or weight and those with signs or symptoms of severe malaria will be referred to the nearest health facility, as they would normally do during their course of work as CHWs delivering iCCM. CHWs also addressed any other health related questions or concerns raised by the household that fell within their iCCM mandate during the visit. Information on fever and fever care provided by the CHW was recorded in their paper-based iCCM register.

ProCCM was only delivered in study arm 1.

### Stock out mitigation

Measures will be taken to ensure there is sufficient supply of malaria commodities to address the increased demand potentially generated by the proactive visits and to mitigate general stock outs. These measures were:

The supply of commodities held at the district level was increased from four to six months of expected use to allow for surplus that may be needed for the proactive intervention in the study district.

The district health management teams (DHMT) and in-charges of PHUs were engaged to facilitate release of at least a month worth of stock to HTR CHWs at the start of the study (*i.e.* a starter commodity pack was provided to CHWs at study trainings).

An SMS based system for HTR CHWs to alert the study team if they are low on supplies was established. Study implementers coordinated with the national medical supplies agency to ensure the study team has access to the commodities needed to ensure resupply, and coordinated transportation of commodities from the central medical stores in Freetown to the selected DHMTs, and coordinated transportation from the DHMTs to the health facilities and/or communities as needed. These interventions were implemented in Arms 1 and 2 of the trial.

### Optimize quality and delivery of SBCC messages

To optimize the quality and delivery of SBCC messages, the national messaging guide was reviewed against the most recent malaria behavior survey to identify any key care-seeking or CHW utilization messages that were missing from the national guidelines and training materials or that required tailoring for the study district. SBCC training materials and job aids delivered in the study district were reviewed for consistency with these updated guidelines. HTR CHWs received training on the SBCC messages to be delivered, with an emphasis on areas where there were previously gaps in the messaging guide or training materials. These messages were implemented only in Arm 1 and 2.

### Arm 3: Routine implementation (control)

No changes will be made to iCCM, SBCC activities, or stocking of HTR CHWs in study arm 3 the control arm. HTR CHWs had already received a 24-day training to prepare them to provide promotional health messages and deliver iCCM services. Per the national strategy, HTR CHWs are expected to visit households in their catchment area once every two months to promote positive health behaviors around hygiene, reproductive health, nutrition, and malaria prevention. HTR CHWs are also expected to screen and treat symptomatic household members for pneumonia, diarrhea, and malaria. HTR CHWs also maintain paper-based registers that are brought to the PHU once a month for entry into the community health information system. At this time, HTR CHWs are intended to be provided with a resupply of commodities. Once a month, HTR CHWs are also supposed to receive supervision from trained peer supervisors.

### Study location

The study was conducted in Pujehun district, where an active CHW network that is already providing iCCM existed. Pujehun District had 104 health facilities, with a wide geographical distribution and diverse healthcare workforce. At the time of trial implementation more than 200 CHWs were in HTR areas or Pujehun district.

**Figure 1.**
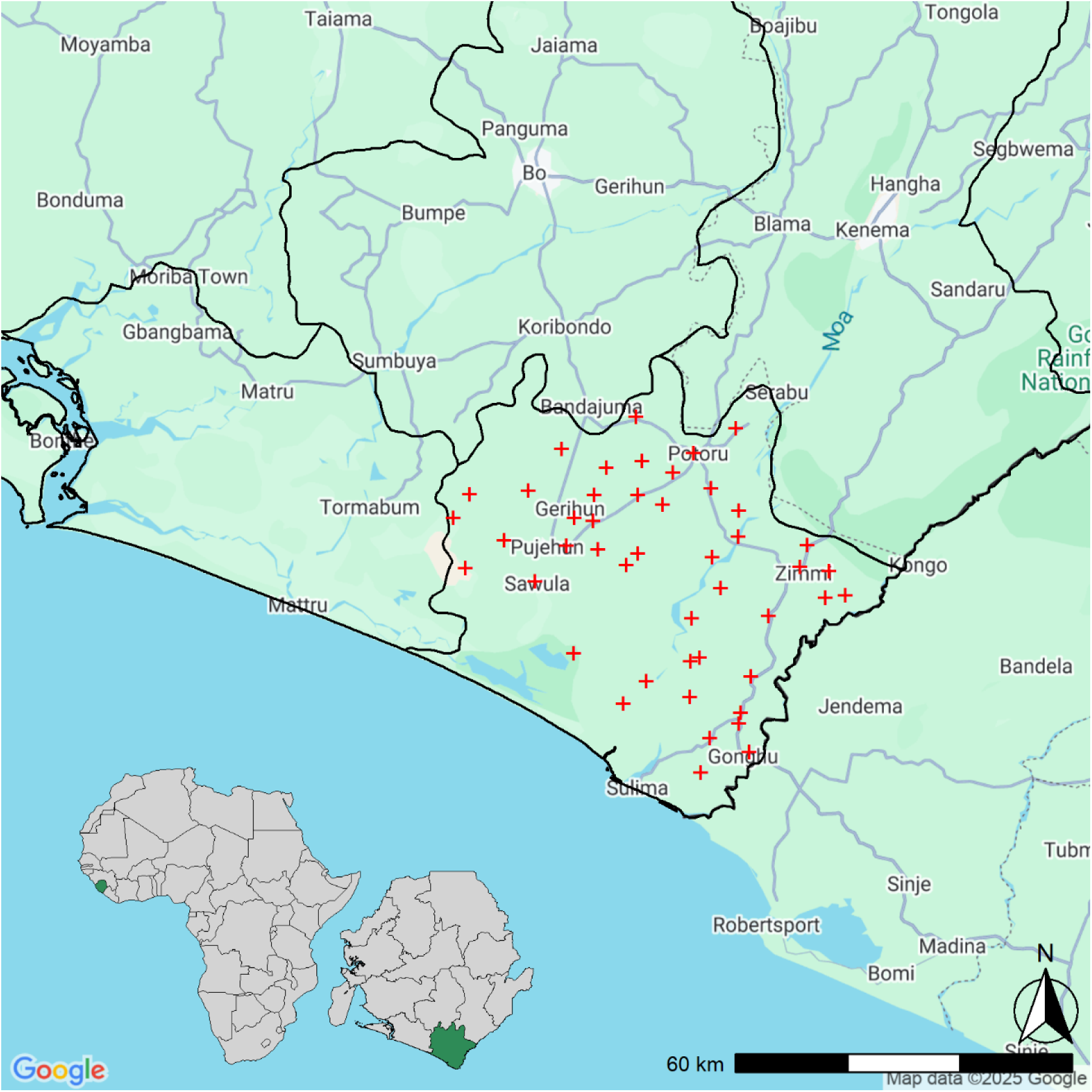
Map of Study Site. Red crosses represent PHCU facilities enrolled in the study.

## Outcomes

The primary-outcome measure for the trial was the proportion of children aged 0-59 months who had a history of fever within the previous 14 days who sought treatment or advice from any provider in the formal care sector on the day of or the day after symptom onset. Fever was defined as any current fever or any history of fever, reported by the caregiver or representative, having occurred within the previous 14 days. For those children who were both age-eligible and had a positive report of fever history, questions were asked to the caregiver as to whether or not care/treatment or advice was sought for the fever. If care/treatment or advice was sought, the timing and source of the first care/treatment or advice seeking attempt in relation to the onset of the febrile symptoms was ascertained. Prompt care seeking was then defined as seeking care on the same day or the day after symptom onset, excluding where care was solely sought from a church, mosque, drug shop/market stall, drug peddler, traditional healer or any combination of these non-formal providers.

### Sample Size

The study was powered to detect changes in the primary outcome, prompt care-seeking. Sample size calculations were conducted using R 4.2.2 (R Core Team (2022). A bespoke MonteCarlo simulation algorithm was developed to simulate a three-armed controlled trial with a binary outcome assuming a logistic random effects data generating model. We assumed a coefficient of variation k ∼ 0.3 and use the Bonferroni correction for multiple testing given the trials three arms require two-tests to determine differences between arms. This results in a *p*-value of <0.025 being required in a two-sided test to control type-I error at the 5% level. The study was expected to have >80% power to detect an increase in early care-seeking from 40% to 60%, with at least 10 children reporting fever in the past two weeks identified in each cluster (assuming 15 clusters per arm (or a total of 45 clusters)).

### Randomization

#### Sequence Generation

Baseline data was used alongside supplemental information about the study clusters to conduct restricted randomization and allocate study clusters to one of the three study arms in a 1:1:1 ratio.

After creation of a cluster level dataset containing summary measures of restriction variables, 70,000 proposed allocation sequences were generated using a random sampling algorithm in R, these allocation sequences were then checked against seven criteria for balance based on levels of baseline prompt care seeking, the average distance of a person in the baseline survey from their PHU, the population size of each cluster (in terms of HHs), and the total number of CHW and the number of communities with no CHW in the cluster. Only 1,106 sequences met all seven criteria. These sequences were then tested for validity, meaning that each cluster must be independently assigned to a study arm, and as such no two clusters should always appear in the same arm or separate arms, and that the frequencies of any pair of clusters appearing together should be similar across all pairs of clusters. The 1,106 sequences generated met these criteria for validity. The study arms were designated as A, B and C at this time and were not assigned to specific interventions.

#### Allocation Concealment Mechanism

Once validity of the list of allocation sequences was established, the trial statistician selected one allocation at random from the list using a sampling algorithm and pseudo random number generator. The assignment of the three groups to specific study arms was done by taking all six possible permutations for assignment of three arms to three groups (*e.g.* A = 1, B = 2, C = 3 or A = 2, B = 1, C = 3 etc…) to the integers one through six by the trial statistician. Subsequently, the head of the national malaria control program of Sierra Leone, who was unaware of any of the allocation sequences, the final cluster selections, or the letter assignment to clusters, rolled a fair six-sided die to choose the arm assignment.

#### Implementation

Training was implemented by study staff in country according to the SOPs developed by the trial team. Implementation of the study interventions was not blinded to the CHW or to the household participants, there was also not formal blinding for data collectors, however, data collectors were not informed about the arm assignment status of the communities where surveys were conducted. The primary analysis script and analysis was conducted by an analyst who was blinded to the arm assignment for the main primary outcome analysis.

#### Analysis

Trial balance was assessed based on the baseline data after randomization as well as through comparisons of background data collected during the endline survey.

As the trial has three arms, the Bonferroni correction for multiple testing was used for the primary analysis, and a *p*-value of <0.025 was required in a two-sided test to control type-I error at the 5% level across both tests.

The primary unadjusted analysis was conducted on the intention to treat analysis population without adjustment for any anticipated confounding variables. A multi-level (variance compartments model) constructed on a generalized linear model framework with a bernoulli likelihood and a logit link function with random intercepts for each study cluster and two indicator variables (one each for arms 1 (ProCCM) and 2 (Stock Support only)) was used to estimate the odds ratio for care seeking based on study arm, as well as to estimate confidence intervals and *p*-values.

#### Adjusted and sensitivity analyses

Adjusted analyses were carried out on the primary outcome to determine whether the estimate of the treatment-effect was changed by the inclusion of additional covariables. Analysis of any effect modifiers were carried out by interacting the covariable with the underlying treatment arm. Arms 2 and 3 were pooled to serve as a control arm for ProCCM per the study SAP. Additionally ideational factors were assed based on responses in the endline survey and were tested for differences across arms similarly to those used for the primary outcome. The time to care seeking was compared between arms using a Kaplan-Meier approach, and the analyses were also repeated on two separate per protocol analysis populations, one in which clusters in the intervention arm were only included when CHWs in the intervention arm completed at least 60% of the anticipated households visits during the intervention period, and a second in which clusters were only included if the CHWs completed at least as many HH visits as expected during the intervention period.

## Results

Across the 45 study clusters, 236 HTR communities containing 7,720 households were enumerated, of which 5,578 were considered eligible for sampling at the time of the first endline survey. For that survey 3,124 households were sampled for screening and 3,069 households with 3,116 children under the age of five were ultimately screened. In these households, enrollment and consent for the parent or caretaker of 393 under five children with fever histories in the previous 14 days, representing 379 individual households, were obtained and the primary study outcome (prompt treatment seeking) was ascertained for 391 children.

**Figure 2.**
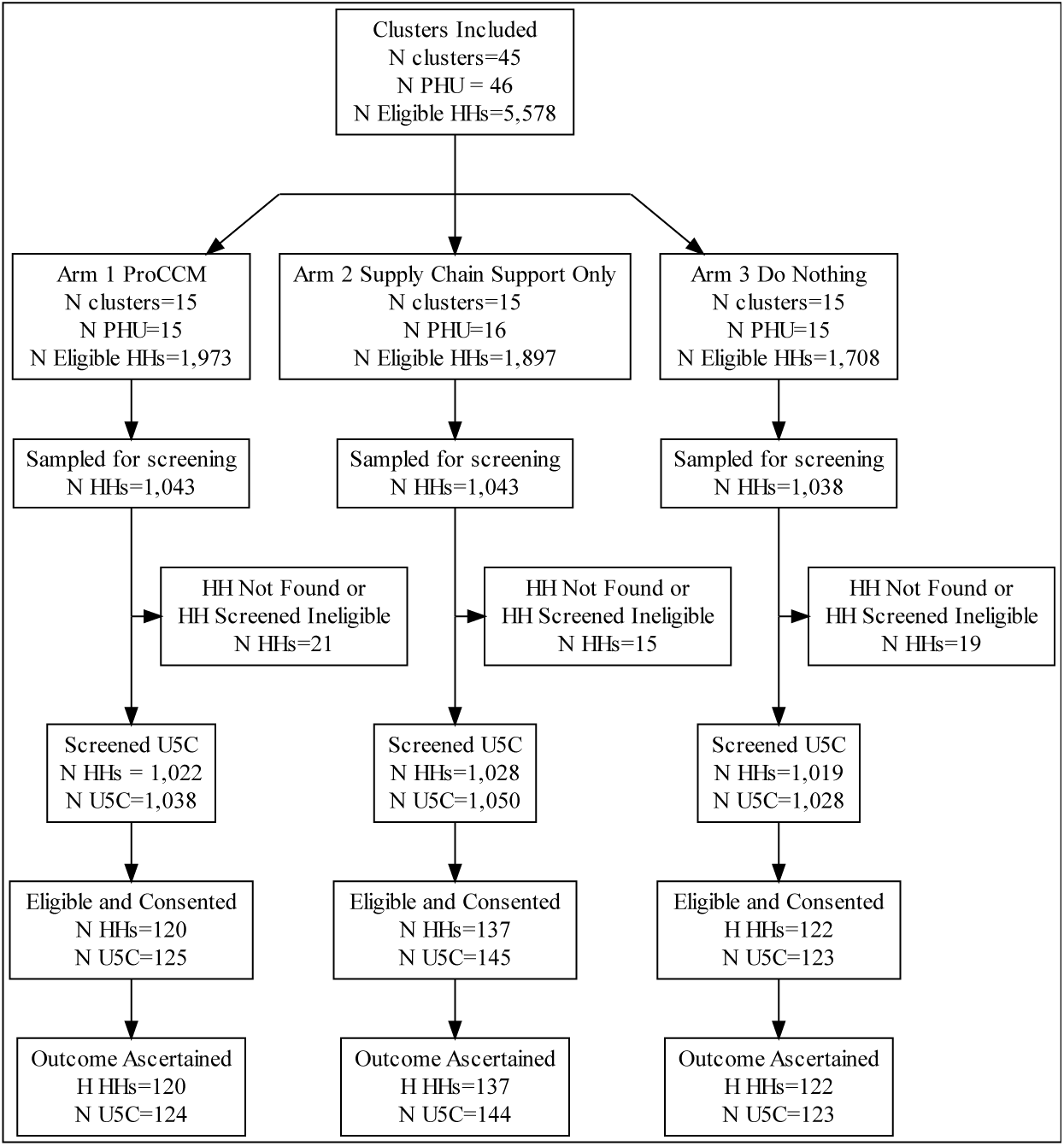
Study CONSORT diagram

Baseline data showed balance across study arms on the primary outcome, with prompt treatment seeking for fever in the formal sector being sought by 55% of children across the three study arms. Some HTR communities lacked dedicated CHWs and were thus served by CHW from nearby communities, but the number of such communities was small (n=4) and they were balanced across study arms with two such communities in the main intervention arm and only one in each of the other two study arms. At the first endline survey the population of these HTR communities had a generally low education level, with more than 50% of the population of household heads or children’s mothers having no formal education. The population was also generally poor and lived in substandard dwellings with approximately 70% of households having earthen floors and more than half of the population lacking access to any kind of toilet or sanitation facility at the household level. Access to malaria prevention was high and more than 90% of households reported that children had slept under an ITN the night before the survey.

The rollout of the first malaria vaccine had begun in Pujehun district and households reported that more than 20% of the children in the study households had received at least one dose of the malaria vaccine.

**Table 1.** Participant Characteristics.

| **Baseline Survey** | **ProCCM** | **Supply Chain** | **Routine Implementation** |
| --- | --- | --- | --- |
| Prompt Care seeking | 54.0% | 55.9% | 55.0% |
| Mean Distance from PHU to HH (km) | 4.2 | 4.0 | 3.8 |
| Mean Number of HH per Cluster | 182 | 174 | 149 |
| Total Number of HTR CHWs | 40 | 38 | 41 |
| Number of Clusters with No HTR CHW at Start of Intervention | 2 | 1 | 1 |
| **Endline Survey 1** | **ProCCM**<br>**N = 120<sup>1</sup>** | **Supply Chain**<br>**N = 137<sup>1</sup>** | **Routine Implementation**<br>**N = 122<sup>1</sup>** |
| **Head of household's education level** |  |  |  |
| None | 76 (63%) | 94 (69%) | 79 (65%) |
| Any (or DK) | 44 (37%) | 43<br>(31%) | 43 (35%) |
| **Household head religion** |  |  |  |
| Christian or Prefer not to say | 5 (4.2%) | 4 (3%) | 1 (0.8%) |
| Muslim | 115 (96%) | 133<br>(97%) | 121 (99%) |
| **Type of floor material for the house** |  |  |  |
| Earth/sand | 80 (67%) | 100<br>(73%) | 87 (71%) |
| Cement | 40 (33%) | 37<br>(27%) | 35 (29%) |
| **Toilet facility for the household** |  |  |  |
| Pit latrine/Septic/Sewer | 61 (51%) | 55<br>(40%) | 56 (46%) |
| Open defecation | 59 (49%) | 82<br>(60%) | 66 (54%) |
| **Primary source of drinking water** |  |  |  |
| Piped or tap water | 34 (28%) | 33<br>(24%) | 29 (24%) |
| Protected well | 48 (40%) | 31<br>(23%) | 33 (27%) |
| Unprotected | 38 (32%) | 73<br>(53%) | 60 (49%) |
| **Mean age of caretaker (sd)** | 34 (12) | 35 (12) | 37 (13) |
| **Gender: Female** | 77 (62%) | 89<br>(62%) | 68 (55%) |
| **Child's mother's education level** |  |  |  |
| None | 66 (53%) | 83<br>(58%) | 67 (54%) |
| Any or Don't Know | 58 (47%) | 61<br>(42%) | 56 (46%) |
| **Slept under net last night.** | 120 (97%) | 136<br>(94%) | 118 (96%) |
| **Received malaria vaccine** | 26 (21%) | 43<br>(30%) | 32 (26%) |

Study interventions were carried out, including household visiting and stock out prevention in study arms 1 and 2. In addition to providing support for stockout prevention, stock out reporting systems were put into place to enable rapid reporting and responses to stockouts of ACT or RDT during the ProCCM implementation and subsequent follow-up period. Reports of stockouts of ACT and RDT were similar across both Arms 1 and 2 of the study. RDT stock outs were reported more frequently in the ProCCM arm (Arm 1) than in the Stock out support only arm (Arm 2) (∼13% of CHW reporting periods vs. ∼9%, and 28% of PHU reporting periods vs. 13%; OR 0.36 95% CI 0.12 – 0.98; *p* = 0.041). ACT stock outs were reported more frequently in the stock out support only arm (Arm 2) than in the ProCCM arm (Arm 1) (∼38% of CHW reporting periods vs. ∼28%, and 47% of PHU reporting periods vs. 43%; OR 1.26 95% CI 0.41 – 4.03; *p* = 0.671). ProCCM visits were conducted in all ProCCM (Arm 1) study clusters and the number of completed HH visits was tracked as compared to the expected number of visits based on the enumerated number of households in each HTR community. The proportion of expected household visits completed ranged from 0.48 to 3.94, in the largest and smallest communities, respectively. The number of expected visits or more was completed in 11 of 15 PHUs (73%) and in only one PHU were fewer than 60% of the expected total household visits completed.

**Table 2.** Study outcomes.

| Arm | N (%) Any Care | N (% of those who sought care) Sought care in formal sector | N (%) Prompt Care in formal sector | N Total |
| --- | --- | --- | --- | --- |
| Arm 1 – ProCCM | 121 (98%) | 120 (99%) | 93 (75%) | 124 |
| Arm 2 - Supply Chain Support Only | 129 (90%) | 121 (94%) | 79 (55%) | 144 |
| Arm 3 – Routine Implementation | 119 (97%) | 119 (100%) | 78 (63%) | 123 |
| Pooled Control | 248 (93%) | 240 (97%) | 157 (59%) | 267 |
| Total | 369 (94%) | 360 (98%) | 250 (64%) | 391 |

The endline survey was conducted between September 20^th^, 2024 and October 3^rd^, 2024. It measured treatment seeking outcomes and other associated individual household and community factors among 391 children less than five years of age whose caretaker reported had a history of fever in the previous fourteen days. Of these children, 64% (250/391) sought care promptly (the day or day after symptom onset) at an appropriate facility (*e.g.* in the formal care sector). Nearly all children were taken for care and advice (94%) and only a small fraction (9/391, ∼2% sought care purely from informal sources such as drug shops or traditional healers). Many children (110/360 31%) were not taken for care or advice to a formal sector facility until the second day after symptom onset (or later) leading to delays in receipt of diagnosis and therapy and the relatively low levels of prompt care seeking seen overall. The proportion of children promptly seeking care was higher in the ProCCM arm (75%) than in either of the two control arms in the study (Arm 2 supply chain support 55% and Arm 3 standard practice / no intervention 63%).

In the main unadjusted ITT analysis, the odds of promptly seeking care were 1.8 times higher (OR 1.84 95% CI 0.94 – 3.82; *p* = 0.077) in the ProCCM arm (Arm 1) than in the Standard Care arm (Arm 3) corresponding to an increase in prompt care seeking of 12 percentage points compared to standard care and 20 percentage points compared to the supply chain only support arm (Arm 2). Supply chain support only (Arm 2) was not meaningfully different than the business as usual, standard care arm (Arm 3) 55% vs. 63% prompt care seeking (OR 0.70 95% CI 0.36 – 1.35; *p* = 0.264). Several additional outcome analyses were planned per the SAP, including pooling the two control arms if differences were not considered statistically different.

This pooled analysis resulted in increased effect size for ProCCM (Arm 1) relative to the pooled control arms (OR 2.24 95% CI 1.23 – 4.10; *p* = 0.009) corresponding to an increase of 16 percentage points in prompt care seeking (59% - 75%). Adjusted analyses on the ITT analytic population were also conducted, which showed significant associations of the outcome with baseline cluster level prompt care seeking. In the pooled analysis, being further than the median distance to the PHU within a cluster significantly increased the effect of ProCCM on care seeking (OR 2.78 95% CI 1.03 - 7.85; *p* = 0.0471). Children in clusters that had higher baseline care seeking also experienced higher care seeking at endline, though here there was no sign of meaningful effect modification. A final adjusted model was selected by AIC and included only adjustment for baseline prompt treatment seeking. This model gave results with similar effect sizes and improved precision in both the three arm analysis and the pooled control formulations. In the three-arm analysis, ProCCM (Arm 1) increased the odds of care seeking promptly by about 1.9 times (OR 1.94 95% CI 1.07 – 3.72; *p* = 0.029), while in the pooled control model it increased the odds of care seeking by about 2.3 times (OR 2.34 95% CI 1.38 – 4.22; *p* = 0.002).

Per-protocol analysis showed similar results, with slightly larger effect sizes and smaller *p*-values. Following the strictest adherence definition (only including clusters that completed 100% or more of the expected household visits, the odds of prompt care seeking was increased by ProCCM (OR 2.69 95% CI 1.28 – 6.24; *p* = 0.0104).

While care seeking overall was nearly universal, as was care seeking in the formal sector, many children experienced delays in care seeking past the day of symptom onset and the following day. Kaplan-Meier analysis showed that the time to care seeking in the formal sector (*p* = 0.034) was significantly longer in the two pooled control arms as compared to the ProCCM arm, indicating that the effect of the intervention in this setting may have been mainly mediated by bringing children into formal care more quickly when febrile, as compared to increasing the frequency at which children sought care overall.

**Figure 3.**
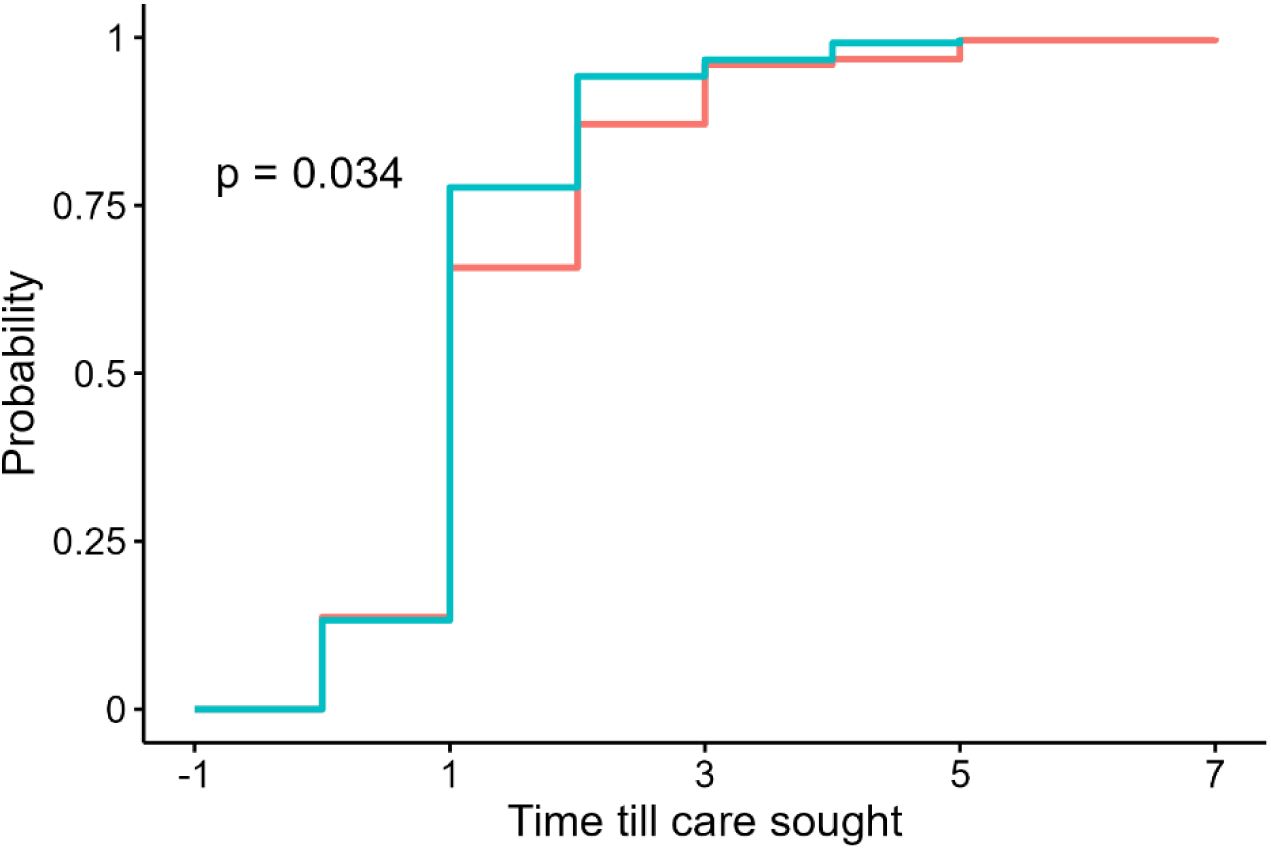
Kaplan Meier survival curves for time to care seeking. Blue line represents ProCCM arm, red line represents pooled control arms.

**Table 3.**
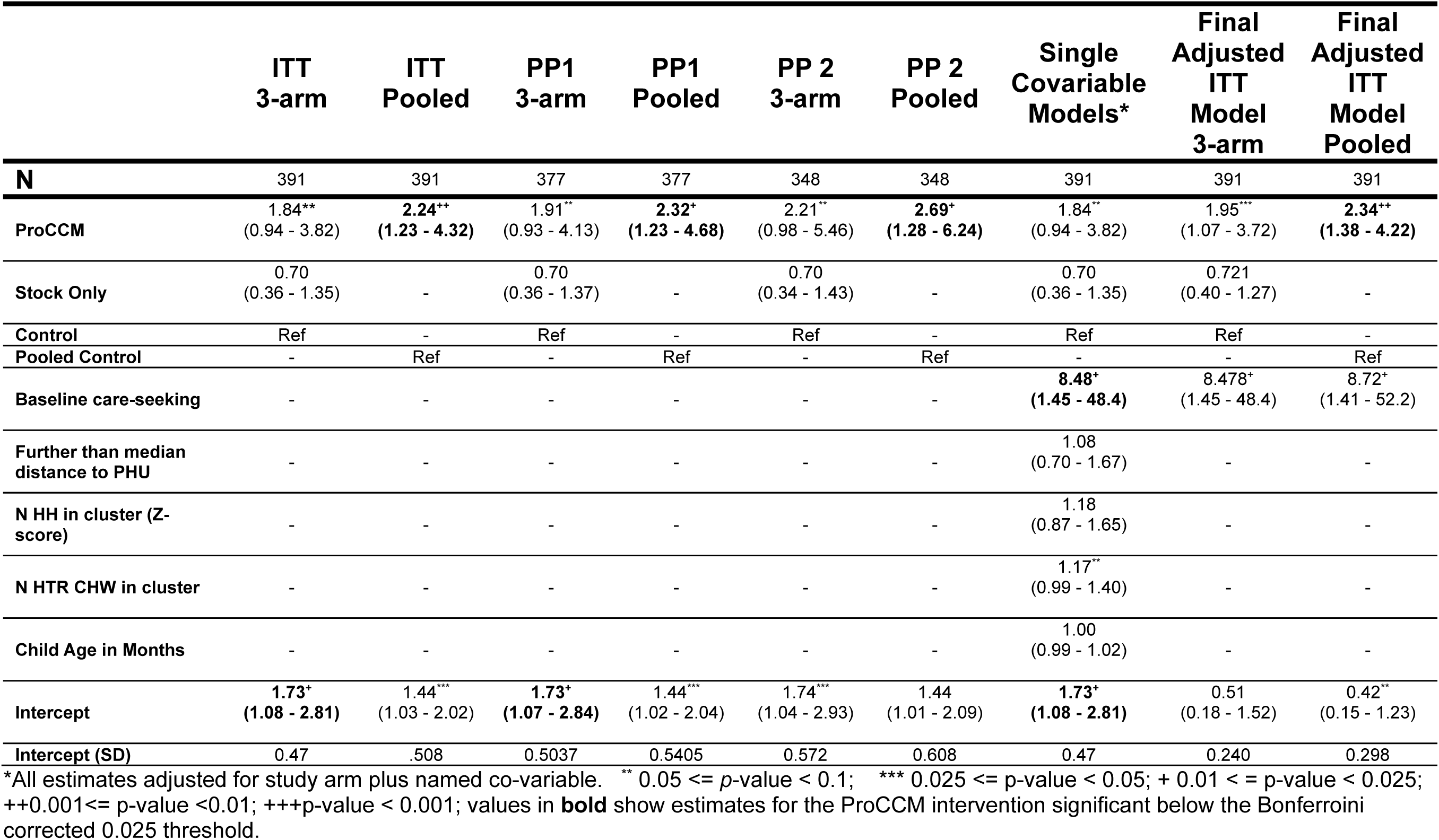
Primary and Secondary Outcome Analyses.

Underlying the hypothesis that ProCCM can improve care-seeking is the presumption that the effect of the intervention is mediated by improving caregivers perceptions of the availability and quality of care being-provided by CHW. To examine the role of the ProCCM intervention we also examined whether the intervention meaningfully improved participants’ perceptions of the quality and availability of care from CHWs. In general, perceptions of CHW based diagnosis and treatment availability and quality were higher in those participants exposed to the ProCCM (Arm 1) intervention compared to the standard care arm (Arm 3) and the SBCC and Stock out support arm (Arm 2) though these only rose to statistically significant differences compared to Arm 3 in the case of availability of medicines and knowledge of treatment in all ages.

**Table 4.** Perceptions of CHW by caregivers and other survey respondents.

| Characteristic | ProCCM<br>N = 120 (HH) | Supply Chain<br>N = 137 (HH) | Routine Imp. (Ref)<br>N = 122 (HH) |
| --- | --- | --- | --- |
| **CHWs treat their patients with respect** |  |  |  |
| Agree *n* (%) | 114 (95%) | 120 (88%) | 112 (92%) |
| OR (95% CI) | 2.11 (0.56 - 9.46) | 0.61 (0.18 - 2.00) | Ref |
| *p*-value | 0.279 | 0.390 | - |
| **CHWs always have medicine to treat malaria** |  |  |  |
| Agree *n* (%) | 80 (67%) | 71 (52%) | 47 (39%) |
| OR (95% CI) | 3.63 (1.58 - 8.99) | 1.74 (0.77 - 4.09) | Ref |
| *p*-value | 0.002 | 0.174 | - |
| **CHWs always have blood test kits for malaria** |  |  |  |
| Agree *n* (%) | 90 (75%) | 85 (62%) | 75 (61%) |
| OR (95% CI) | 1.76 (0.81 - 3.82) | 0.95 (0.45 - 2.00) | Ref |
| *p*-value | 0.153 | 0.898 | - |
| **CHWs know how to treat malaria in all ages** |  |  |  |
| Agree *n* (%) | 92 (77%) | 81 (59%) | 77 (63%) |
| OR (95% CI) | 1.99 (1.03 - 3.84) | 0.79 (0.43 - 1.44) | Ref |
| *p*-value | 0.040 | 0.441 | - |
| **CHWs make me pay for medication to treat malaria** |  |  |  |
| Disagree *n* (%) | 98 (82%) | 117 (85%) | 109 (89%) |
| OR (95% CI) | 0.45 (0.12 - 1.80) | 0.83 (0.20 - 3.40) | Ref |
| *p*-value | 0.260 | 0.796 | - |
| **When children in your community have fever, what proportion are taken to a CHW to get tested for malaria?** |  |  |  |
| All children *n* (%) | 63 (53%) | 56 (41%) | 52 (43%) |
| OR (95% CI) | 1.64 (0.60 - 4.50) | 0.85 (0.31 - 2.32) | Ref |
| *p*-value | 0.334 | 0.746 | - |

No harms were reported or identified during the trial, which was of home visiting, and thus was not considered to be a high risk of expected harms occurring aside from loss of privacy and confidentiality. While unexpected harms could have occurred during the course of trial, none were reported. The trial was stopped early due to the dismantling of the United States Agency for International Development, and consequently the complete and abrupt removal of funding for this study. Consequently, the final survey round could not be completed. As the intervention period was already complete, and the outcome assessments were conducted using cross sectional methods, no participants were considered “on-study” at the time of interruption.

## Discussion

This community randomized three-armed trial was designed to demonstrate that a short period of ProCCM could improve and sustain changes in treatment seeking behavior for fever in a population in a predominately rural area of Sierra Leone. This is important given that it has been shown that malaria community case management (mCCM) can be effective at reducing the malaria burden, but ancillary benefits such as sustained changes in care seeking behaviors have not been robustly assessed and therefore limited information is available to guide deployment strategy for iCCM programs (*e.g.* whether to use passive post based iCCM or to use an outreach based strategy such as ProCCM). This study demonstrates that ancillary benefits may indeed exist in the form of changes in care-seeking behavior which extend beyond the direct benefits of extending CCM models to rural communities with limited access to health facilities.

Unfortunately, due to the abrupt interruption of work on of the President’s Malaria Initiative through stop work orders and disruptions caused by the dismantlement of USAID, the trial was interrupted, preventing to completion of the last round of outcome assessment and thereby determining the longer-term sustainability of these changes. The major limitation of the trial is thus that while it gives definitive evidence that ProCCM can improve care seeking beyond the simple availability of CCM with a robust supply chain, it is not clear how long the improved care seeking behaviour will persist after the ProCCM intervention is stopped.

The trial relies on a self-reported outcome, care seeking behavior, which is subject to recall bias. Indeed, studies have documented that patient recall of many parts of the clinical encounter in similar populations may be highly unreliable. However, the general aspects of care seeking (whether, what type of care, and how soon care was sought after symptoms began) have been shown to be reliable. The specific phraseology and approach to assess treatment seeking used here have been widely and consistently used to assess these behaviours throughout sub-Saharan Africa. Further, uniform assessment in a well randomized study reduces the chances that any recall bias would be differential between arms.

As care seeking was nearly universal (as was use of the formal sector) in this population it was not possible to improve care seeking for this population by improving the fraction of children taken for care overall. Nor was it possible to shift care from the informal sector to the formal sector, since the fraction seeking care in informal settings alone was vanishingly small.

As such, the only avenue to improve care seeking practice in this setting was by reducing the time from symptom onset to care seeking, which clearly occurred in the context of the ProCCM intervention. This avenue for improvement of care would likely be available in any setting in which prompt care seeking in the formal sector levels were below optimal, since even an absence of care seeking behaviour entirely could be viewed as an extended delay rather than a choice not to seek care at all. The high level of care seeking in the formal sector may, however, pose some challenges in generalizing this study result to areas in which the overall care seeking in the formal sector is lower. Changes in perception caused by ProCCM might not be able to overcome access barriers such as high costs of care or extreme travel to access care. In this setting, however, participants in the hardest to reach settings benefited more than those in slightly less hard to reach settings and the overall context was one of very limited care access. We believe that the findings in this study are thus likely to be translatable to other settings in sub-Saharan Africa, including remote rural locations.

Indeed, several studies of ProCCM have indicated that improvements in general care seeking might occur after this intervention, though none so robustly designed and conducted as this study.

The changes in ideational factors surrounding the availability and quality of care observed in the ProCCM arm (Arm 1) give a strong indication as to a plausible mechanism through which a short-term application of ProCCM could act causally to change behavior. The house visiting shortened the time to care seeking rather than improving identification of prevalent symptomatic infections (which it may also do). These changes are likely mediated by changing the perceptions of caregivers about the quality, availability and importance of taking children promptly for care in this setting, and do not seem to result from simply extending CCM and enduring stock of drugs and RDTs since they were not apparent in Arm 2. Rather it appears that the outreach occurring during household visits seems to result in changed perceptions leading to more prompt care seeking behaviour in the context of ProCCM.

Delays in care seeking for malaria illness (along with other febrile Illnesses) can result in serious consequences including progression to severe malaria and potentially death.

Thus, increases in prompt care seeking offer the potential to lead to increased child survival, and other long-term benefits known to result from malaria burden reductions. This study did not directly measure such benefits, however. Additionally, it is also likely that implementing ProCCM comes at an additional cost compared to iCCM alone.

Whether the additional benefits derived from ProCCM are likely to be cost-effective relative to other interventions for preventing morbidity and mortality from malaria is an open question and will require further study.

## Conclusion

Proactive Community Case Management implemented over a short period (∼2 months), can improve care seeking behavior in communities, especially those where access barriers such as travel time to formal care may be highest. Whether such benefits can be sustained over time or whether the additional cost of ProCCM as compared to CCM is more cost-effective than other interventions to reduce malaria morbidity and mortality remain open questions.

## Other Information

Registration: Clinicaltrials.gov ID: NCT06395207

Protocol and SAP: Included as Supplementary Information.

Funding: USAID/PMI Funding through PATH/PMI Insights supported the original study and data collection. No funding was provided for analysis or manuscript preparation.

## Key Roles and Contact Information

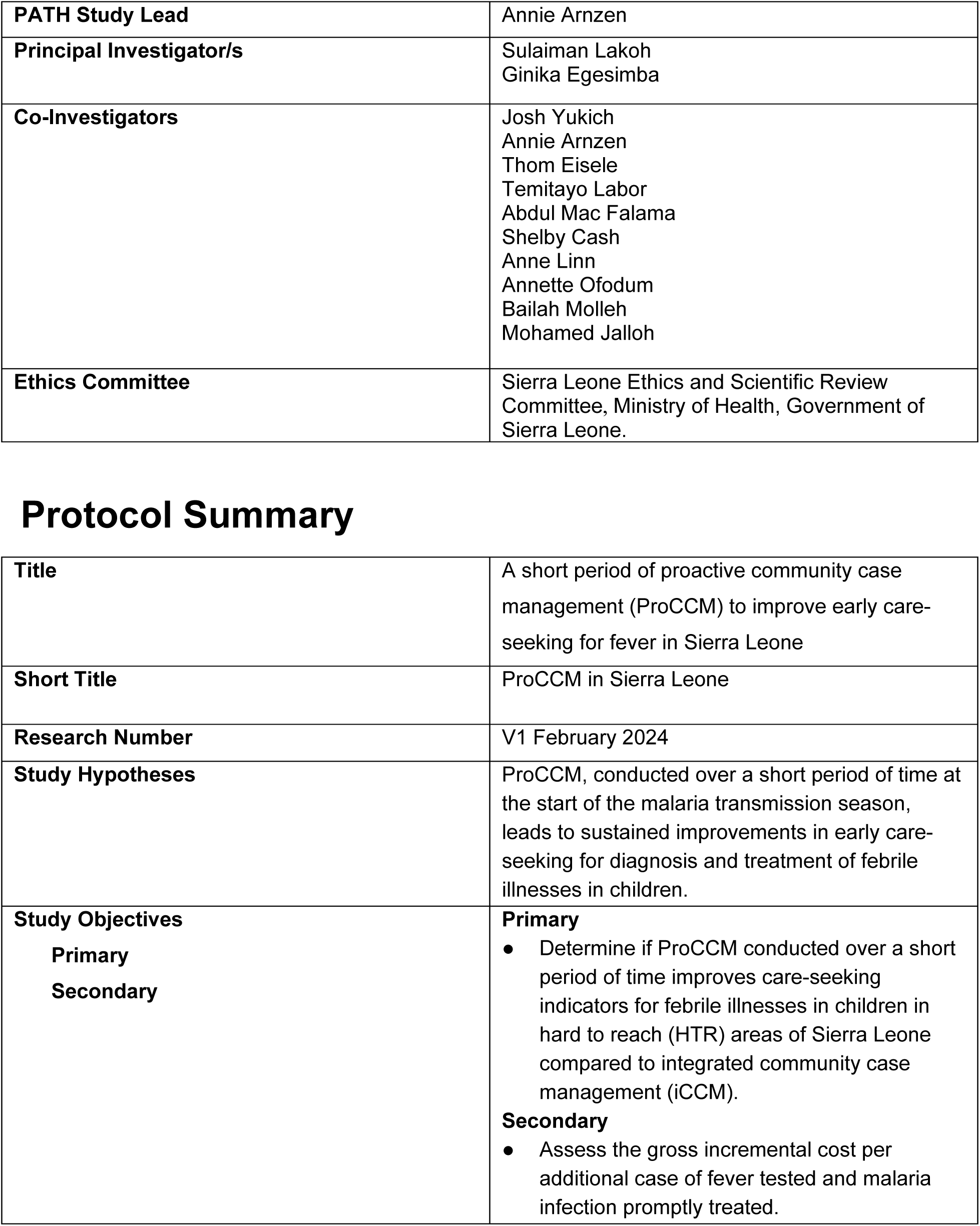
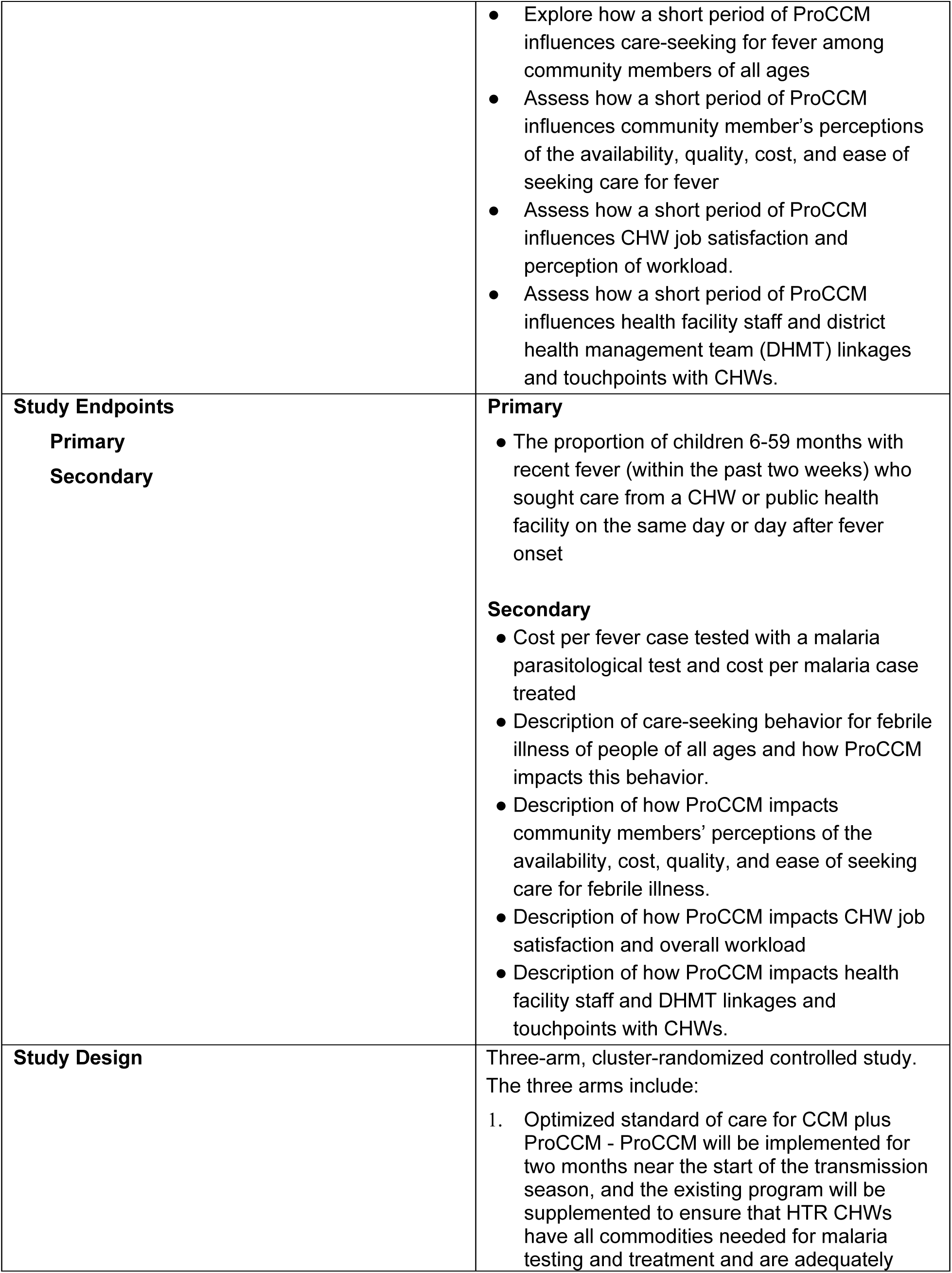
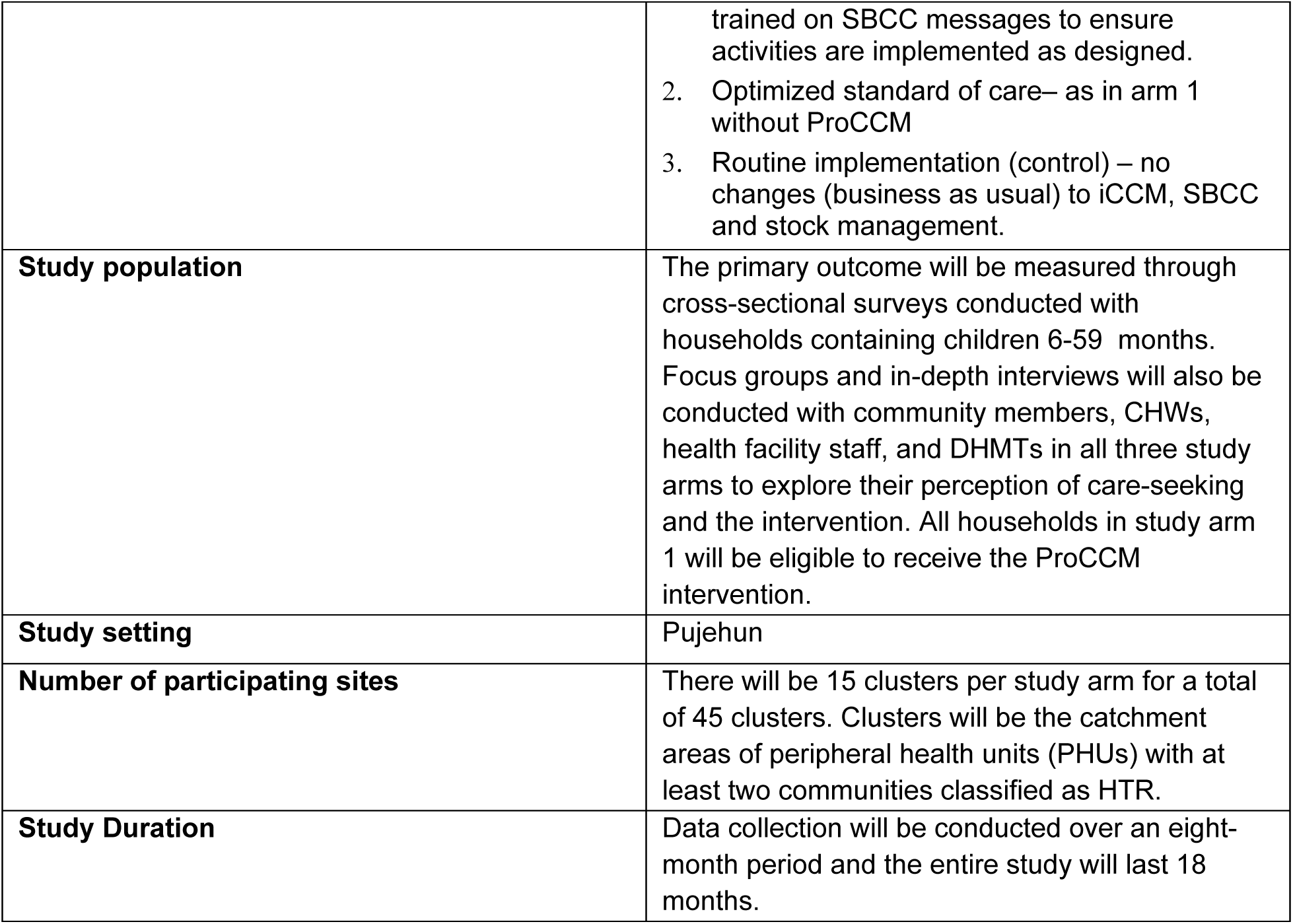

|  |  |
| --- | --- |
| **PATH Study Lead** | Annie Arnzen |
| **Principal Investigator/s** | Sulaiman Lakoh<br>Ginika Egesimba |
| **Co-Investigators** | Josh Yukich<br>Annie Arnzen<br>Thom Eisele<br>Temitayo Labor<br>Abdul Mac Falama<br>Shelby Cash<br>Anne Linn<br>Annette Ofodum<br>Bailah Molleh<br>Mohamed Jalloh |
| **Ethics Committee** | Sierra Leone Ethics and Scientific Review Committee, Ministry of Health, Government of Sierra Leone. |
| Protocol Summary |  |
| **Title** | A short period of proactive community case management (ProCCM) to improve early care-seeking for fever in Sierra Leone |
| **Short Title** | ProCCM in Sierra Leone |
| **Research Number** | V1 February 2024 |
| **Study Hypotheses** | ProCCM, conducted over a short period of time at the start of the malaria transmission season, leads to sustained improvements in early care-seeking for diagnosis and treatment of febrile illnesses in children. |
| **Study Objectives**<br>**Primary**<br>**Secondary** | **Primary**<br>Determine if ProCCM conducted over a short period of time improves care-seeking indicators for febrile illnesses in children in hard to reach (HTR) areas of Sierra Leone compared to integrated community case management (iCCM).<br>**Secondary**<br>Assess the gross incremental cost per additional case of fever tested and malaria infection promptly treated. |
|  | • Explore how a short period of ProCCM influences care-seeking for fever among community members of all ages<br>• Assess how a short period of ProCCM influences community member's perceptions of the availability, quality, cost, and ease of seeking care for fever<br>• Assess how a short period of ProCCM influences CHW job satisfaction and perception of workload.<br>• Assess how a short period of ProCCM influences health facility staff and district health management team (DHMT) linkages and touchpoints with CHWs. |
| **Study Endpoints**<br>**Primary**<br>**Secondary** | **Primary**<br>• The proportion of children 6-59 months with recent fever (within the past two weeks) who sought care from a CHW or public health facility on the same day or day after fever onset<br>**Secondary**<br>• Cost per fever case tested with a malaria parasitological test and cost per malaria case treated<br>• Description of care-seeking behavior for febrile illness of people of all ages and how ProCCM impacts this behavior.<br>• Description of how ProCCM impacts community members' perceptions of the availability, cost, quality, and ease of seeking care for febrile illness.<br>• Description of how ProCCM impacts CHW job satisfaction and overall workload<br>• Description of how ProCCM impacts health facility staff and DHMT linkages and touchpoints with CHWs. |
| **Study Design** | Three-arm, cluster-randomized controlled study. The three arms include:<br>1. Optimized standard of care for CCM plus ProCCM - ProCCM will be implemented for two months near the start of the transmission season, and the existing program will be supplemented to ensure that HTR CHWs have all commodities needed for malaria testing and treatment and are adequately |
|  | trained on SBCC messages to ensure activities are implemented as designed.<br>2. Optimized standard of care– as in arm 1 without ProCCM<br>3. Routine implementation (control) – no changes (business as usual) to iCCM, SBCC and stock management. |
| **Study population** | The primary outcome will be measured through cross-sectional surveys conducted with households containing children 6-59 months. Focus groups and in-depth interviews will also be conducted with community members, CHWs, health facility staff, and DHMTs in all three study arms to explore their perception of care-seeking and the intervention. All households in study arm 1 will be eligible to receive the ProCCM intervention. |
| **Study setting** | Pujehun |
| **Number of participating sites** | There will be 15 clusters per study arm for a total of 45 clusters. Clusters will be the catchment areas of peripheral health units (PHUs) with at least two communities classified as HTR. |
| **Study Duration** | Data collection will be conducted over an eight-month period and the entire study will last 18 months. |

## Acronyms

CHW community health worker

CCM community case management

DHMT ETR district health management team easy-to-reach

HTR hard-to-reach

iCCM integrated community case management

ITN insecticide treated net

NMCP national malaria control program

PHU peripheral health units

ProCCM proactive community case management

SBCC social behavior change communications

## 1. Background and rationale for the study

### Background

Malaria case management that includes testing and treatment of people experiencing signs and symptoms of illness is an essential strategy in the fight against malaria in many countries in Africa. The utilization and impact of these services is largely dependent on people seeking care when they or their children have fever. In many malaria-affected countries, trends in care-seeking have not seen the same marked improvements that have been observed in other intervention coverage indicators^1^. The barriers to care-seeking are well documented including the cost of health services, travel time and distance, and negative perceptions regarding the quality of care available^2–4^.

In recent years, many countries have introduced and scaled up community case management (CCM) of malaria (and, in many settings, integrated management of childhood illness or iCCM, which includes malaria, pneumonia, and diarrhea) through community health workers (CHWs). The availability of malaria services at the community level through CHWs addresses geographic barriers to care and increases access to life-saving services^5^. However, the utilization of CHWs varies widely by context^1,6^. The low utilization of CHWs is impacted by many factors. To date, CHWs programs have not consistently addressed the known barriers to care such as user fees for services, inadequate CHW supervision, and provision only for patients who sought care from a fixed site^7^. Stock out of essential supplies is another important factor that influences the acceptability and confidence in CHWs^8^. Preliminary findings from a literature review conducted by Breakthrough Action found that drivers of community utilization of CHWs include trust, satisfaction, and perception of CHW skills and knowledge (Breakthrough ACTION presentation on CHW Literature Review, unpublished). The review also found that sending CHWs to houses more frequently has a positive impact on community utilization of CHW services.

To reduce the gap in care-seeking and address some of the known barriers to seeking care and utilizing CHW services, proactive case finding strategies have been implemented in some countries to increase the prompt treatment of uncomplicated malaria and prevent such cases from developing into severe disease. One such strategy is proactive community case management (ProCCM). In this intervention, CHWs visit households in their communities to proactively offer a defined package of services, such as CCM for malaria or iCCM. In models of this intervention that focus on malaria, CHWs screen for persons with fever and offer malaria diagnostic and other assessment services, as well as malaria treatment, health communication, and referral services. Proactive community case management is implemented in addition to the CHWs’ routine activities providing passive CCM or iCCM in community settings. Studies of ProCCM have been conducted in a number of different countries and settings, with recent studies in Senegal^9,10^, Madagascar^5^, Mali^11,12^, and Zambia (results unpublished). In these studies, the frequency and duration of the proactive intervention has ranged from weekly to biweekly visits, with coverage either during the malaria transmission season only or year round.

ProCCM has been shown to be well accepted by the community, feasible to implement, and increases the number of patients treated by CHWs^9,10^. In some settings, the implementation of ProCCM has been linked to improvements in early care-seeking for fever^9^. In Senegal, a case study of scaling up ProCCM found that the number of malaria cases detected and treated by CHWs increased during periods of passive work conducted in between proactive rounds^10^. The largest increase in care-seeking during passive periods was seen during the first year of implementation, and continued, albeit at lower levels, during year two and three of the intervention scale-up^10^. A study in Mali also found notable improvements in prompt care-seeking for febrile illness in the context of a ProCCM program with the rate of early effective antimalarial treatment of children under five increasing from 14.7 percent to 35.3 percent^11^.

However, a systematic review conducted by Whidden et al found that while many studies of proactive case-finding home visits by CHWs increased access to care and reduced child mortality, there was no significant impact on care-seeking behavior^7^. The certainty of the evidence available to date on the relationship between ProCCM and care-seeking is low and requires further research with rigorous study design.

### Context in Sierra Leone

Malaria remains a significant public health problem in Sierra Leone and is the leading cause of illness and death among children under five years of age. According to the 2021 Malaria Indicator Survey, malaria prevalence is 22 percent among children aged 6-59 months^13^. Malaria transmission in Sierra Leone is perennial with seasonal peaks occurring during the early rainy season in May and again in October, at the end of the rainy season. The primary malaria prevention efforts include universal coverage with insecticide treated nets (ITNs), intermittent preventive treatment in pregnancy (IPTp), and perennial malaria chemoprevention (PMC).

Health services are delivered through a three-tiered system including peripheral health units (PHUs), district/secondary hospitals, and national/regional referral hospitals^14^. There are 1428 functional health facilities, including 65 hospitals, 258 community health centers (CHCs), 433 community health posts (CHPs), and 672 maternal and child health posts (MCHPs). The government of Sierra Leone is the main healthcare provider. Other healthcare providers include faith-based local and international non-governmental organizations (NGOs) and the private sector. Sierra Leone has a large private health sector, but the exact size is unknown.

The community health program serves as an extension of the PHUs. Sierra Leone first introduced a CHW program in 2012, which was then scaled nationwide in 2017. The program was reintroduced in 2022 after a two-year hiatus during which time the CHW policy was updated to establish an integrated approach to scale up malaria diagnosis and include an integrated package of services at the community level^15^. The approximately 8,154 current CHWs provide an integrated package of services. CHWs in easy-to-reach (ETR) areas serve a catchment population of 500-1,000 in 100 to 170 households located between three to five kilometers from the nearest PHU. ETR CHWs are responsible for providing health counseling and making referrals to the PHU. Hard-to-reach (HTR) CHWs serve a catchment population of 300 to 350 in 50 to 60 households in areas that are more than five kilometers from the nearest PHU or between 3km and 5km from the nearest PHU in areas with difficult terrain^15^. CHWs in ETR and HTR areas are expected to proactively visit households in their community once every two months to provide SBCC messages, and preventive and promotional services. However, SBCC messages are not consistently delivered in all areas and CHWs are not trained on all key messages included in the national message guide for malaria.

In addition to health prevention and promotional services, HTR CHWs are trained and supplied to provide expanded iCCM services, including diagnosis and treatment of malaria, diarrhea, and pneumonia.

However, HTR CHW stock outs are a known barrier to providing reliable testing and treatment services. An assessment conducted by the USAID Global Health Supply Chain Program in Sierra Leone in 2023 found that over 40 percent of health facilities were stocked out of malaria commodities needed to supply CHWs at the time of the assessment.

The Sierra Leone 2021 Malaria Indicator Survey found that advice or treatment was sought for 75 percent of children who had fever within the preceding two weeks, however only 40 percent sought care on the same or next day following fever onset^13^. Despite the availability of CHWs, only 7 percent of caregivers who sought care for a child with recent fever reported seeking care from a CHW, with the majority seeking care from a government health facility (68 percent)^13^. Enabling factors that have been shown to influence care-seeking in Sierra Leone include self-efficacy, awareness of CHWs as a resource, positive attitude about the benefit of prompt care-seeking, and open dialogue with their spouse about malaria^16^. In a recent malaria behavior survey (MBS), 87% of respondents indicated CHWs know how to treat malaria in children^16^. There is an opportunity to build upon this confidence and build awareness of CHWs. Improving trends in care-seeking has been identified as a priority by the National Malaria Control Program (NMCP) and donors.

### Rationale for the study

Despite the critical importance of timely identification and treatment of malaria, many families do not promptly seek care for febrile illness for a variety of reasons. In Sierra Leone, ProCCM household visits may serve to improve the visibility of CHWs to the community, increase the community’s level of awareness of CHW capabilities, strengthen the community’s trust in CHWs and, therefore, increase the community’s utilization of CHWs even during periods when ProCCM household visits are not occurring (figure 1).

**Figure 1:**
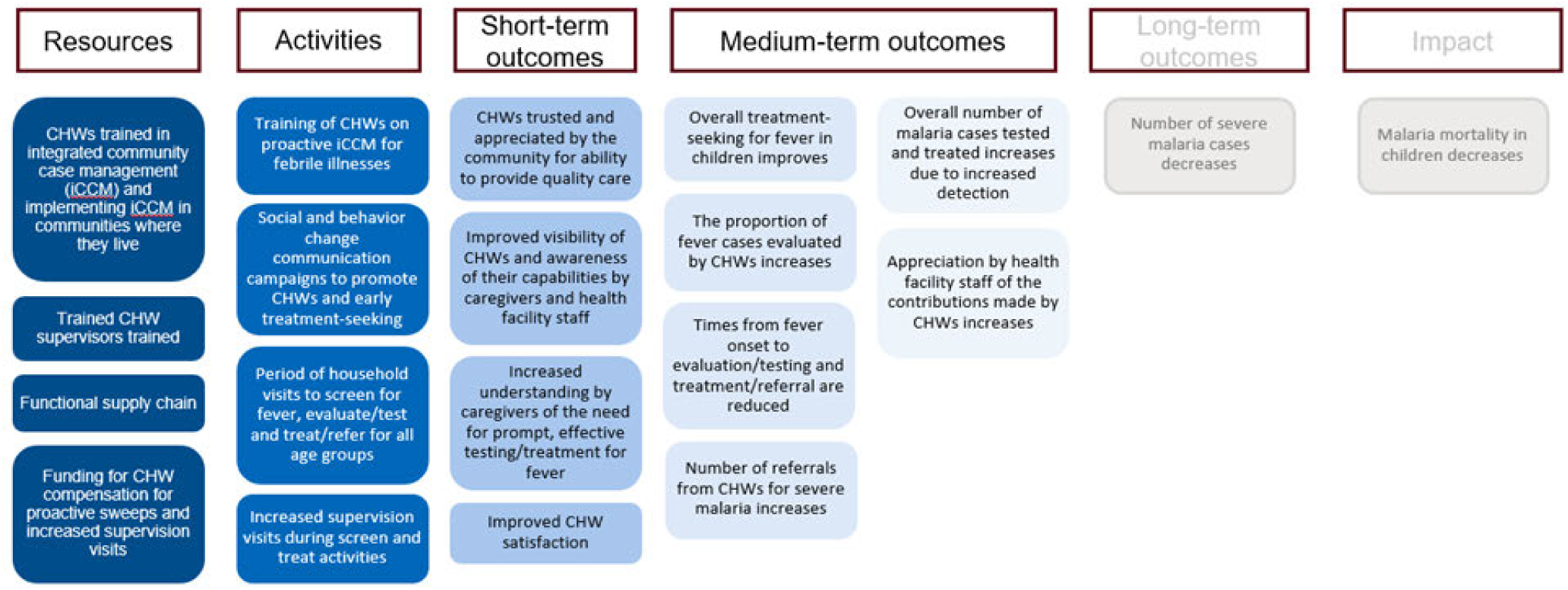
Theory of change for the impact of a short period of ProCCM on early care-seeking^1^ ^1^ Many of the studies conducted to date have focused on the impact of proCCM on malaria transmission as measured by changes in parasite prevalence, with mixed results.

While ProCCM has been shown to be a promising intervention for increasing care-seeking behavior, regular rounds of ProCCM household visits have typically lasted at least throughout the entire transmission season and often throughout the year, making it an expensive intervention to maintain. Further, there is insufficient evidence on the sustained impact of ProCCM on care-seeking behavior.

This study seeks to determine whether, among communities living in HTR areas of Sierra Leone, a short period of ProCCM (specifically, four household visits in two months) conducted at the start of a peak malaria transmission season can improve early care-seeking for fever and whether these improvements are sustained for the duration of the malaria transmission season.

## 2. Study hypothesis, objectives, and endpoints

The primary aim of this study is to assess the impact of a short period of ProCCM on care-seeking with the hypothesis that ProCCM conducted over a short period of time leads to sustained improvements in early care-seeking for febrile illnesses in children in HTR areas of Sierra Leone compared to iCCM.

### 2.1. Primary and secondary objectives

The primary objective of this study is to:

- Determine if ProCCM conducted over a short period of time improves care-seeking indicators and utilization of CHWs for febrile illnesses in children in HTR areas of Sierra Leone compared to iCCM.

The secondary objectives are to:

- Assess the gross incremental cost per additional case of fever tested and malaria infection promptly treated. Explore how a short period of ProCCM influences care-seeking for fever among community members of all ages
- Assess how a short period of ProCCM influences community member perceptions of the availability, cost, quality and ease of seeking care for febrile compared to iCCM.
- Assess how a short period of ProCCM influences CHW job satisfaction and impact on overall workload.
- Assess how a short period of ProCCM influences health facility staff and DHMT linkages and touchpoints with CHWs.

### Study endpoints

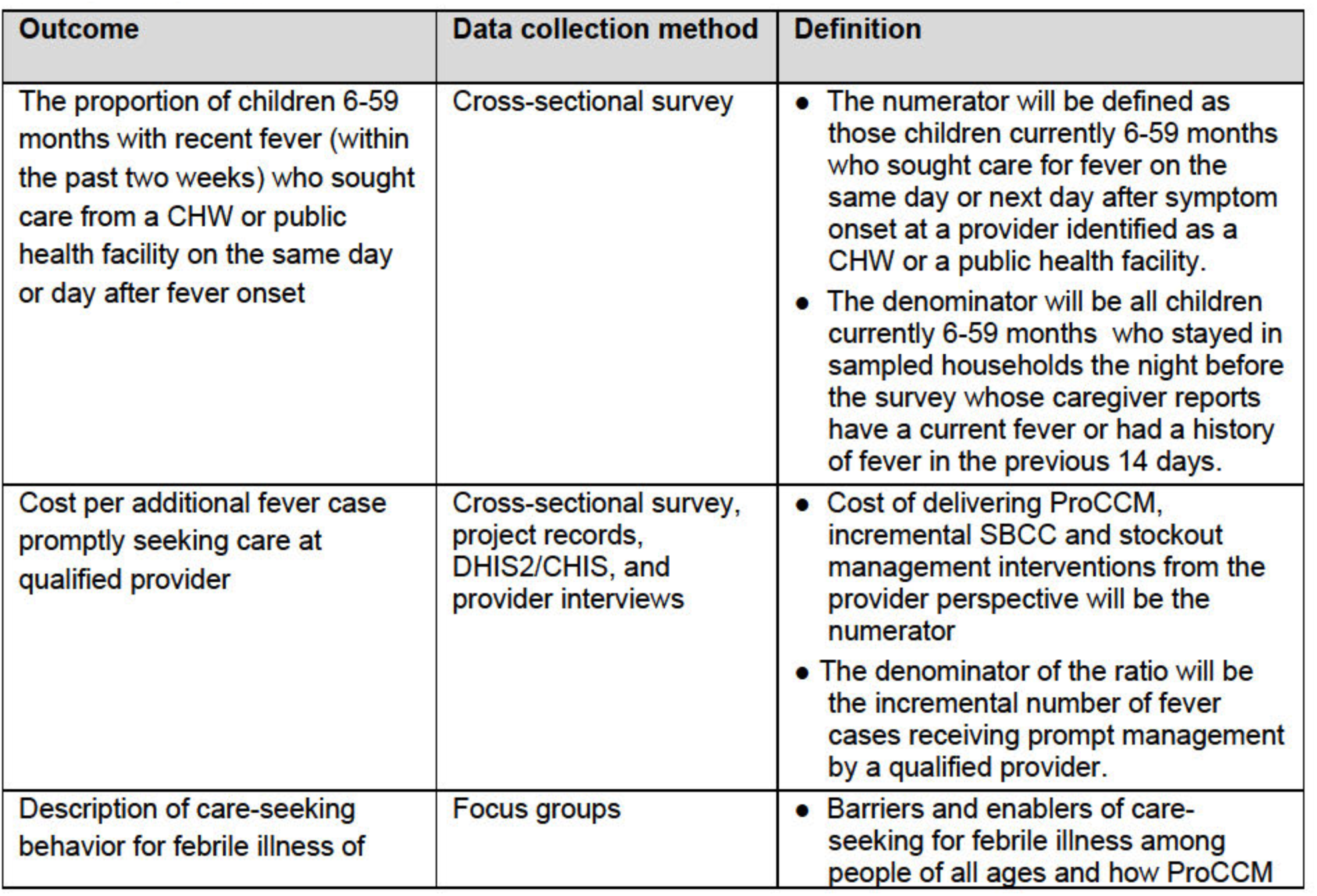
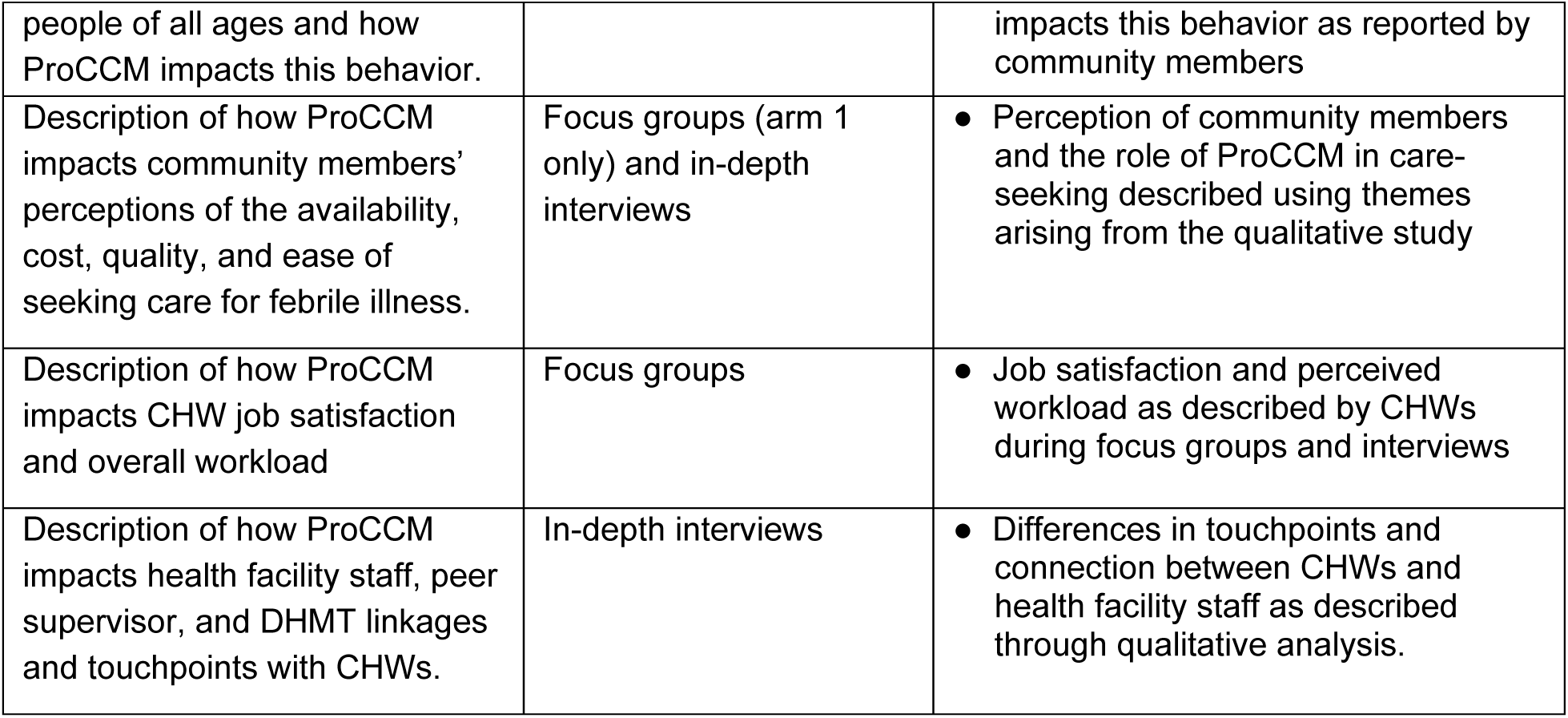

| Outcome | Data collection method | Definition |
| --- | --- | --- |
| The proportion of children 6-59 months with recent fever (within the past two weeks) who sought care from a CHW or public health facility on the same day or day after fever onset | Cross-sectional survey | • The numerator will be defined as those children currently 6-59 months who sought care for fever on the same day or next day after symptom onset at a provider identified as a CHW or a public health facility.<br>• The denominator will be all children currently 6-59 months who stayed in sampled households the night before the survey whose caregiver reports have a current fever or had a history of fever in the previous 14 days. |
| Cost per additional fever case promptly seeking care at qualified provider | Cross-sectional survey, project records, DHIS2/CHIS, and provider interviews | • Cost of delivering ProCCM, incremental SBCC and stockout management interventions from the provider perspective will be the numerator<br>• The denominator of the ratio will be the incremental number of fever cases receiving prompt management by a qualified provider. |
| Description of care-seeking behavior for febrile illness of | Focus groups | • Barriers and enablers of care-seeking for febrile illness among people of all ages and how ProCCM |
| people of all ages and how ProCCM impacts this behavior. |  | impacts this behavior as reported by community members |
| Description of how ProCCM impacts community members' perceptions of the availability, cost, quality, and ease of seeking care for febrile illness. | Focus groups (arm 1 only) and in-depth interviews | • Perception of community members and the role of ProCCM in care-seeking described using themes arising from the qualitative study |
| Description of how ProCCM impacts CHW job satisfaction and overall workload | Focus groups | • Job satisfaction and perceived workload as described by CHWs during focus groups and interviews |
| Description of how ProCCM impacts health facility staff, peer supervisor, and DHMT linkages and touchpoints with CHWs. | In-depth interviews | • Differences in touchpoints and connection between CHWs and health facility staff as described through qualitative analysis. |

## 3. Methods

### 3.1. Study design

This is a three-arm, cluster-randomized controlled study to be conducted in Pujeun district in Sierra Leone. Clusters will be the catchment areas of peripheral health units (PHUs) with at least two communities classified as HTR and where there is at least one CHW trained and active in iCCM. The study arms will consist of the following interventions, described in detail in section 3.2:

- ptimized standard of care for CCM plus ProCCM - ProCCM will be implemented for two months near the start of the high transmission season, and the existing program will be supplemented to mitigate stock outs of malaria commodities among HTR CHWs and ensure HTR CHWs are adequately trained on SBCC messages to ensure activities are implemented as designed.
- ptimized standard of care – as in arm 1 without ProCCM
- implementation (control) – no changes (business as usual) to iCCM, SBCC and stock management.

Changes in care seeking will be assessed through three cross-sectional surveys. A baseline survey will be conducted prior to intervention implementation followed by a second survey at or near the end of the ProCCM intervention period and a final survey at the end of the malaria transmission season. Cost of the intervention will be measured following standard ingredients approach based micro-costing strategies and used to assess the cost and cost-effectiveness of the intervention to improve care seeking behavior.

Interviews or focus groups will be conducted with individuals who chose to promptly seek treatment and those who did not in response to a fever in all three study arms to explore how ProCCM impacts community members’ perceptions of the cost, ease, quality, and benefits of seeking care. Interviews will also be conducted with HTR CHWs in all three study arms to explore job satisfaction and perceived workload and with health workers around their touchpoints and linkages with CHWs. Lastly, focus groups will be held with community members of all ages to explore their care-seeking behavior for febrile illness.

### 3.2. Interventions

In all three study arms, HTR CHWs will continue to provide the standard package of iCCM (passive case detection) plus services as described in the national CHW strategy. HTR CHWs serve a catchment population of 300 to 350 people (50 to 60 households) that is more than five kilometers from the nearest health facility or three to five kilometers in areas with particularly difficult terrain. HTR CHWs will be supervised by peer supervisors who have been trained and are in operation according to the national strategy.

#### 3.2.1. Arm 1: Optimized standard of care for CCM plus ProCCM

##### ProCCM

The ProCCM intervention will be implemented at the beginning of the peak malaria transmission season with an anticipated duration of two months. HTR CHWs and supervisors in study arm 1 will receive additional training, supported by the national CHW hub, on the proactive intervention. CHWs will be expected to visit each household in their catchment area twice per month for a total of four home visits per household. An additional monetary incentive will be provided to HTR CHWs in arm 1 to compensate them for the proactive visits added to their existing scope of work.

During the visits, CHWs will identify household members of all ages complaining of fever or history of fever and record household profile in a paper-based register. People with fever, history of fever in the past 48 hours, or symptoms suggestive of malaria will receive an RDT. CHWs will provide individuals that test malaria positive with a course of antimalarials based on national guidance for dosing based on age and/or weight and those with signs or symptoms of severe malaria will be referred to the nearest health facility. CHWs will also address any other health related questions or concerns raised by the household that fall within their iCCM mandate during the visit. Information on fever and fever care provided by the CHW will be recorded in their paper-based iCCM register.

##### Stock out mitigation

The below measures will be taken to ensure there is sufficient supply of malaria commodities to address the increased demand generated by the proactive visits and to mitigate stock outs.

- Increase from four to six months of supplies held at the district level to allow for surplus that may be needed for the proactive intervention in the study district.
- Engage DHMT and in-charges of PHUs to facilitate release of at least a month worth of stocks to HTR CHWs at the start of the study (i.e. starter pack provided at trainings).
- Establish an SMS based system for HTR CHWs to alert the study team if they are low on supplies. SHS to coordinate with PMI and national medical supplies agency (NMSA) to ensure the study team has access to the commodities needed to make these resupplies (e.g. SHS will coordinate transportation of commodities from the central medical stores in Freetown to the selected DHMTs, and coordinate transportation from the DHMTs to the health facilities and/or communities as needed.

##### Optimize quality and delivery of SBCC messages

To optimize the quality and delivery of SBCC messages, the national messaging guide will first be reviewed against the malaria behavior survey to identify any key care-seeking or CHW utilization messages that are missing from the national guide and training materials or that require tailoring for the study districts. Once the national malaria messaging guide has been updated as needed, the SBCC training materials and job aids delivered in the study district will be reviewed to ensure all key messages are included. HTR CHWs will receive training on the SBCC messages to be delivered, with an emphasis on areas where there were previously gaps in the messaging guide or training materials.

#### 3.2.2. Arm 2: Optimized standard of care

Study arm 2 will be the same as study arm 1 only without the implementation of proactive visits every two weeks for two months.

#### 3.2.3 Arm 3: Routine implementation (control)

No changes will be made to iCCM, SBCC activities, or stocking of HTR CHWs in study arm 3. HTR CHWs have received a 24-day training to prepare them to provide promotional health messages and deliver iCCM services. Per the national strategy, HTR CHWs are expected to visit households in their catchment area once every two months to promote positive health behaviors around hygiene, reproductive health, nutrition, and malaria prevention. HTR CHWs are also expected to screen and treat symptomatic household members for pneumonia, diarrhea, and malaria. HTR CHWs maintain paper-based registers that are brought to the PHU once a month for entry into the community health information system (CHIS). At this time, HTR CHWs are intended to be provided with a resupply of commodities. Once a month, HTR CHWs are also supposed to receive supervision from trained peer supervisors.

##### Limitations

Study clusters will be designed around PHU catchment areas and randomly assigned to study arms. There is a potential for a spillover effect with community members seeking out health services in the intervention arms due to improvements in community case management (e.g. availability of stock).

### 3.3. Study location

The study will be conducted in Pujehun, one of the three districts where PMI is supporting an active CHW network that is already providing iCCM. Study clusters will be designed around HTR areas in the study districts that are currently served by an active CHW network. Pujehun District is situated in Sierra Leone’s Southern Province. The healthcare system addresses prevalent issues such as malaria, respiratory infections, and diarrheal diseases through robust public health campaigns and educational initiatives. Healthcare delivery challenges, including transportation difficulties, communication barriers, and inadequate resources, hinder the overall health situation. Pujehun District has 104 health facilities, with a wide geographical distribution and diverse healthcare workforce, including community health workers. In particular, 234 CHWs are in HTR areas, playing a pivotal role in extending healthcare outreach to these remote regions.

### 3.4. Sampling methods and sample size

#### Sampling and sample size for cross-sectional surveys

The study is powered on changes in the primary outcome, early care-seeking. Sample size calculations were conducted using R 4.2.2 (R Core Team (2022). R: A language and environment for statistical computing. R Foundation for Statistical Computing, Vienna, Austria. URL https://www.R-project.org/.) The approach to sample size calculation involved the use of a bespoke MonteCarlo simulation algorithm which simulated a three-armed controlled trial with a binary outcome assuming a logistic random effects data generating model. We assume a coefficient of variation k ∼ 0.3 and use the Bonferroni correction for multiple testing given the trials three arms require two-tests to determine differences between arms. This results in a p-value of <0.025 being required in a two-sided test to control type-I error at the 5% level. We estimate that the study will have >80% power to detect an increase in early care-seeking from 40% to 60%, with at least 10 children reporting fever in the past two weeks identified in each cluster (assuming 15 clusters per arm (or a total of 45 clusters)).

To find 10 febrile children 6-59 months, we estimate 60 households will need to be screened in each cluster during each survey round, assuming there will be an average of 1 child in that age group per household and that each child has an 11-17% two-week period prevalence of fever. Therefore, 900 households will be visited and screened in each arm for a total of 2,700 households per cross-sectional survey and 8,100 households in total. There will be a potential to interview up to 8,100 caretakers (should every screened household have a child with a history of febrile illness in the previous two weeks) about care-seeking history for fever in their children during the study, but the expected number of interviews is much lower, approximately 1,500. Sample **size** calculations were also checked against the formula for a two-armed trial with a binary outcome provided by Hayes and Moulton (H & **M** 2017), assuming k = 0.3, and 10 individuals per cluster, 15 clusters per arm should provide sufficient power >80% to detect a change from 40% to 60% in early care-seeking at a significance level of alpha = 0.025.

Sampling will be conducted by simple random sampling without replacement from the household list (limited to HH with children expected to be between 6 and 59 months of age at the time of the survey) stratified by survey cluster.

#### Sampling and sample size for interview and focus group discussions

Caregivers will be purposively selected from each of the three study arms with the aim of identifying individuals/households who chose to promptly seek care and those who did not in response to a fever. HTR CHWs and health workers will also be purposively selected with the aim of understanding the impact of ProCCM on CHWs workload and coordination with the health facility following study implementation. A representative from the DHMT team will also be interviewed. In-depth interviews will be conducted with community members, health workers, and the DHMT due to ease of logistics and to allow for open, transparent responses to the interview questions. Focus group discussions (FGDs) will be organized with HTR CHWs when they convene at the PHU for their monthly meeting. Three to five HTR CHWs will be included in FGD depending on the PHU catchment size. Approximately 67 in-depth interviews and 13 focus groups will be conducted across all three study arms to achieve saturation of perspectives and themes. Table 1 below describes the proposed sample size by study arm.

**Table 1:** Proposed sample for interviews and focus group discussions.

|  | Arm 1<br>(ProCCM) | Arm 2<br>(optimized<br>standard of<br>care) | Arm 3<br>(Control) | Total |
| --- | --- | --- | --- | --- |
| Individuals/households that promptly sought care | 8 IDIs | 8 IDIs | 8 IDIs | 24 IDIs |
| Individuals/households that did not promptly seek care | 8 IDIs | 8 IDIs | 8 IDIs | 24 IDIs |
| Individuals of all ages with recent fever | 4 FGDs |  |  |  |
| HTR CHWs | 3 FGD | 3 FGD | 3 FGD | 9 FGD |
| HTR CHW peer supervisors | 3 IDIs | 3 IDIs | 3 IDIs | 9 IDIs |
| PHU in-charge | 3 IDIs | 3 IDIs | 3 IDIs | 9 IDIs |
| DHMTs |  |  |  | 1 IDIs |
| Total: 13 FGDs, 67 IDIs |  |  |  |  |

## 4. Study procedures

### 4.1. Overview

As depicted in figure 2, preparatory activities including community mapping and enumeration, baseline cross-sectional survey, randomization to study arms, community sensitization and engagement, and CHW and data collector trainings will be conducted before launching the study intervention. Following randomization to the three study arms, the study intervention will begin with implementation for two months. Cross-sectional surveys, in addition to the baseline survey, will be collected at two timepoints: 1) immediately after the cessation of the ProCCM intervention to assess changes in care-seeking in the short term due to the ProCCM intervention and 2) significantly after the intervention to measure sustained changes 3-4 months after intervention. Qualitative data collection will be collected towards the end of the transmission season to explore community, HTR CHW, and health facility staff’s perceptions of care-seeking and the influence of ProCCM on these perceptions.

**Figure 2:**
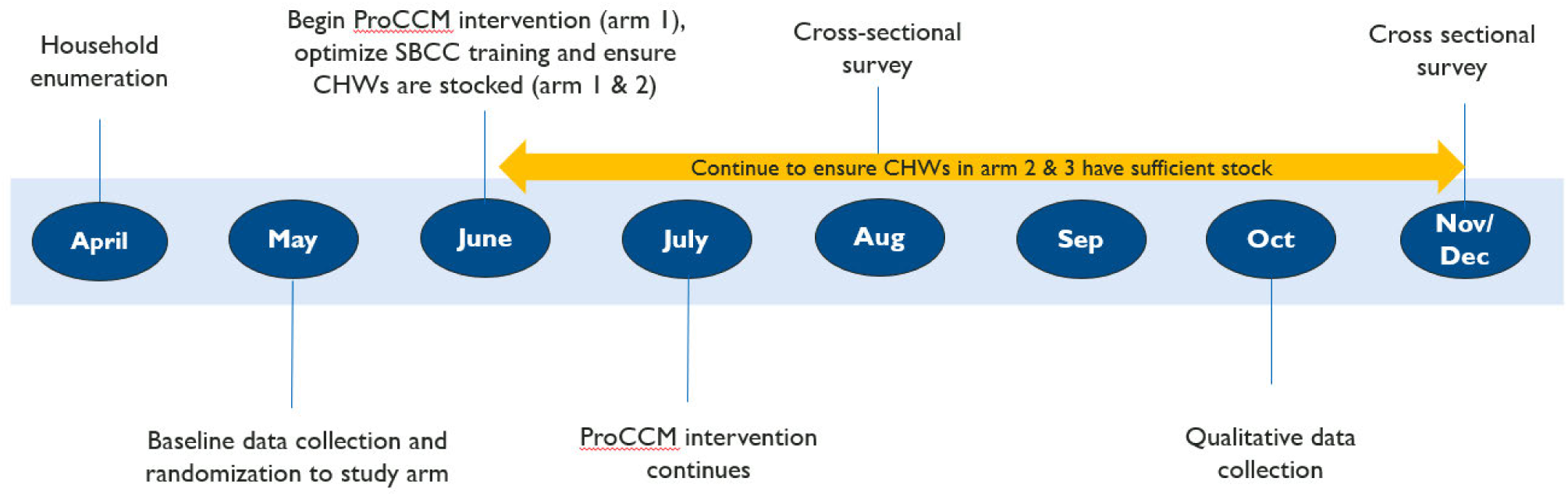
Overview of intervention and data collection timeline

### 4.2. Mapping and enumeration

All households (HHs) that fall within the defined clusters (e.g. HTR CHW catchments within a selected PHU) will comprise the sampling frame for measurement of outcomes, and will be geolocated, mapped, and enumerated.

#### 4.2.1. Mapping of study clusters

Clusters will be established at all HTR CHW locations within a PHU (see figure 4). Prior to baseline data collection, at least 45 clusters will be established across study arms by identifying PHUs and mapping HTR CHWs around PHUs. Cluster boundaries will be established through participatory mapping with HTR CHW and all households within the catchment areas of the HTR CHW in chosen or potential clusters will be enumerated and mapped. Participatory mapping and cluster boundaries may be established using preliminary mapping data from WorldPop or other satellite imagery source to create base maps for initial cluster boundary proposals. Attempts will be made to ensure that clusters are geographically separated to minimize the risk of contamination due to CHWs serving households outside of their established service areas.

**Figure 4:**
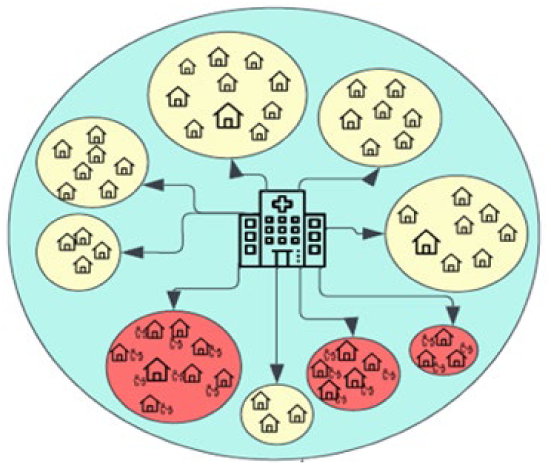
Design of study clusters with HTR CHW locations denoted in red

Clusters that present significant logistical or operational challenges for data collection, and/or that demonstrate at baseline to have no/very little malaria transmission or are outliers with much higher transmission than is typical, will not be included in the final set of study clusters.

#### 4.2.2 Enumeration

Each study cluster will be enumerated (households in the catchments of HTR CHWs) prior to a baseline survey and randomization for all HTR CHW areas located within the cluster (PHU). The enumeration will consist of confirming whether any children under five live in a household, if so documenting the number of children under five years of age living in each household, and taking GPS locations for all households located within the HTR areas of the PHU (cluster).

##### Enumeration eligibility

- Household is inside the catchment of a HTR CHW inside a selected PHU
- Verbal consent from adult respondent to participate in the enumeration.

##### Enumeration procedures

- team of data collectors will work with the HTR CHW from that community to visit all households within the cluster boundary and within the catchment of that HTR CHW. This process will be repeated for all HTR CHW within each selected cluster (PHU).
- arriving at a household, the data collectors will record the GPS coordinates of the household.
- collectors will ask an adult representative if any members of their household are between 6 and 59 months of age and if there are any children between 0 and 6 months of age.

- If there are no children between 0 and 59 months of age, the data collector will record GPS coordinates and the head of household name.
- If there are children between 6 and 59 months of age, the data collector will record GPS coordinates, the head of household name, and the number of children between 6 and 59 months of age.
- If there are children between 0 and 6 months, the data collector will record GPS coordinates, the head of household name, and the number of children between 0 and 6 months of age.

### 4.3 Baseline cross-sectional survey

A random sample of 60 households in each cluster with children between 6 and 59 months of age will be drawn from the enumeration data set. Baseline data collection will employ a structured survey instrument consisting of a screening questionnaire to determine household eligibility (*e.g.* presence of child between 6-59 months of age with a history of fever in the preceding 14 days (annex X), a consent procedure (annex y), and a full caretaker interview for households in which a child with a history of fever is present (annex z). The questionnaire will be essentially identical to that used for the outcome assessment surveys and will be administered to caretakers of children with a history of fever or a knowledgeable adult representative for the child and caretaker. The instrument for the full interviews will capture information about the child, the household, as well as other demographic and socio-economic information including perceptions of care seeking and typical practices for febrile children. The screening visit is expected to take no more than 20 minutes, while the full interview could last up to one hour.

#### Baseline survey eligibility

- Household is located within at HTR CHW catchment area of a selected study cluster
- Adult respondent consents to participate in the baseline survey
- A child between 6 and 59 months of age is a *de jure* resident of the household and has a history of fever within the previous 14 days.

#### Procedures for Baseline Cross Sectional Survey

- Households expected to have a *de jure* resident between the ages of 6 and 59 months will be sampled from all HH enumerated in each study cluster using simple random sampling.
- Sampled households will be visited by the survey team. Information about the survey will be provided to the head of household or his/her representative and a brief screening will be administered. If the household head and/or his/her representative is not available at the time of the first visit, the household will be visited up to two additional times at varying times of day to limit non-response. Sampled households that cannot be contacted within three visits will not be included in the study, and they will not be replaced with another household.
- If the sampled household is considered eligible after the screening questionnaire *(i.e.* there is a child in the household between the ages of 6 and 59 months who has a history of fever within the previous fourteen days and a caretaker or representative capable of answering questions for the HH), then a full consent and interview will be conducted in consenting households.
- A questionnaire will be administered to the caretaker of the child or his/her representative and will include questions about household demographics, housing characteristics, malaria prevention, and fever treatment-seeking behavior (see annex z). The questionnaire will use electronic data capture administered using tablets.

### 4.4 Randomization to study arm

Randomization will be conducted by the trial statistician following the baseline survey. Randomization will be conducted using a restricted randomization approach whereby each of the clusters will be randomly allocated to one of the three study arms (1:1:1) by selection of an acceptable randomization from a large number of proposed randomizations, each of which meets a pre-determined set of restriction criteria (see figure 5). Restriction criteria for each cluster for balancing the study arms will be based on data obtained from the baseline survey and from the local malaria control program. The main restriction criteria will be balance in prompt care-seeking between arms. The results of the randomization will be shared with the public during community sensitization meetings.

**Figure 5.**
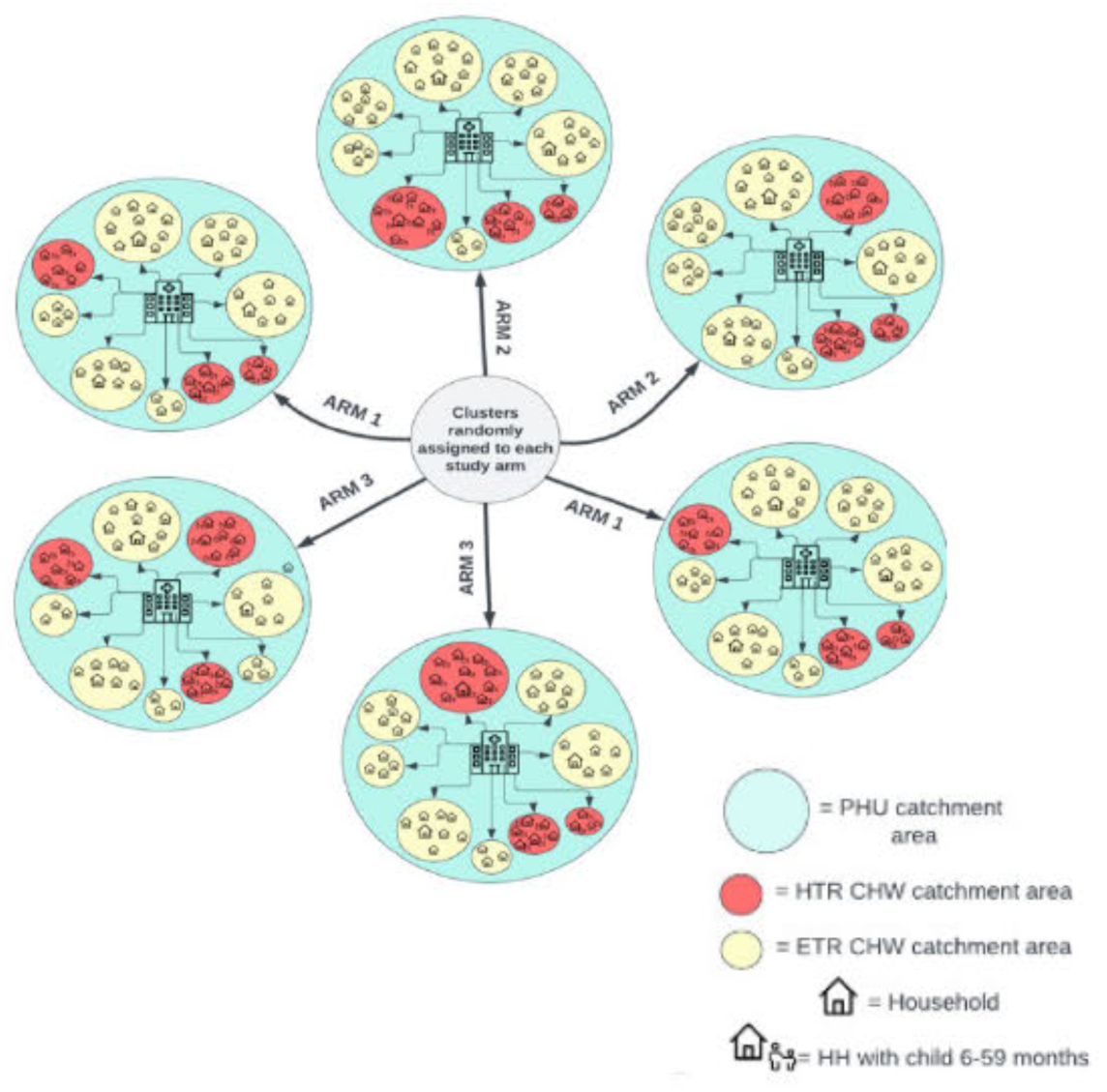
Overview of randomization approach

### 4.5 ProCCM Intervention

HTR CHWs in study arm 1 will receive supplemental training and an additional monetary incentive to increase the frequency of their proactive household visits to two visits per month to each household in their catchment area. The training will focus on the proactive visit schedule and important SBCC messages, as well as a refresher on screening for and treating malaria, given HTR CHWs are already trained on the package of iCCM services. During the proactive visits, HTR CHWs will provide the standard package of iCCM services.

#### Eligibility for ProCCM Intervention

- Household lives within the study cluster
- Head of household or a household member over the age of 18 provides verbal consent for the HTR CHW visit
- All available household members at the time of the HTR CHW visits are eligible to receive services

#### Procedures

- Using their household register as a guide, HTR CHWs in study arm 1 will visit all households in their catchment area two times every month for two months.
- During the household visit, HTR CHWs will screen household members for recent fever and signs and symptoms of malaria. If a household member has had a recent fever or signs and symptoms of malaria, the HTR CHW will perform an RDT. If a household member is RDT positive, the HTR CHW will provide a full course of antimalarials.
- During the proactive visit, the HTR CHW will also assess and treat household members with signs and symptoms of diarrhea and pneumonia. If the HTR CHW does not have the needed supplies, they will refer the symptomatic household member to the nearest health facility.
- HTR CHWs will refer any household members with signs of severe illness to the nearest health facility.
- HTR CHWs will record all activities performed and treatment, if provided, in their paper-based register.
- HTR CHWs will send an SMS to their supervisor if they are running low on any of their commodities.
- HTR CHWs will report to the nearest PHU each month for resupply and to submit their paper based register (HF4 form) for upload to the Community Health Information System (CHIS/DHIS2).

### 4.6 Cross-sectional surveys

Cross-sectional surveys will be repeated at or near the end of the ProCCM intervention period and at the end of the malaria transmission season. The procedures outlined above for the baseline survey will be repeated, but with a new sample of households drawn from the full original enumeration (of households expected to have a child between 6 and 59 months of age) using simple random sampling, stratified by study cluster at each survey round.

### 4.7 In-depth interviews and focus groups

Following intervention implementation, qualitative information will be collected through in-depth interviews or focus group discussions using a semi-structured guide for CHWs, DHMTs, health workers and community members. Qualitative data collection is expected to take two weeks. The eligibility criteria for all individuals to be considered for qualitative data collection is included below.

#### Eligibility by stakeholder group

|  |  |
| --- | --- |
| Community member | • Lives in a study cluster<br>• Reported having a recent fever or family member with recent fever<br>• 18 years or older<br>• Consents to participate in the interview |
| HTR CHW | • Serves as a HTR CHW in a study cluster during the time of the study<br>• 18 years or older<br>• Consents to participate in the FGD |
| HTR CHW Peer Supervisor | • Serves as a HTR CHW peer supervisor in a study cluster during the time of the study<br>• 18 years or older<br>• Consents to participate in the interview |
| PHU In-Charge | • Works at a PHU within a study cluster catchment during the time of the study<br>• 18 years or older<br>• Consents to participate in the interview |
| Senior DHMT official | • Works in the study district at the time of the study<br>• Responsible for coordinating with the CHW focal point on CHW activities<br>• 18 years or older<br>• Consents to participate in the interview |

#### Procedures

- Participants will be oriented to the study and the purpose of the interview and asked to consent to participate (annex xx). Those that provide consent will continue with participation in the discussion.
- Focus groups and interviews will be conducted in a private location and will be closed to outside observers, limited only to consenting participants and the data collectors conducting the interview.
- Data collectors will facilitate the interviews using a semi-structured guide (annex xx).
- FGDs will last approximately 90 minutes and in-depth interviews will last approximately 60 minutes.
- Each FGD and/or interview will be recorded and transcribed for later data analysis.

### 4.7 Cost and cost-effectiveness

Data from project records and interviews with key stakeholders in intervention implementation will be used to construct a description of the intervention implementation as well as to produce estimates of the cost of intervention implementation separately for each cluster and also by the intervention component (SBCC, stock management and ProCCM). Costs will be coded by line item and activity and the cost drivers for each arm (training, stock out mitigation, per diems) will be compared descriptively. These costs will be calculated for one season of implementation. Costs will be primarily collected using the provider perspective and a micro-costing (ingredients) approach will be used for establishment of input quantities and prices to estimate total costs of intervention implementation. Additionally, costs of increased care-seeking from the patient’s perspective will be estimated using individual level data collected in cross-sectional surveys. These data will be combined with intervention costs to estimate the gross costs of the intervention from the societal perspective and supplemented with information from published literature where necessary. The primary outcome will be the gross societal incremental cost per incremental febrile illness case receiving prompt care from a qualified provider. Outcomes will be primarily calculated on a one-year time frame. Sensitivity analysis (one-way, scenario and probabilistic sensitivity analyses (PSA)) will be conducted to examine the effects of various assumptions included in cost and cost-effectiveness models.

## 5. Ethical Considerations

### 5.1. Risks and benefits

There is minimal risk associated with the proposed data collection activities. The main risk associated with the study is the potential threat to an individuals’ privacy and a loss of confidentiality of their data. The privacy of an individual could be compromised during the administration of the survey questionnaire or during focus groups/in-depth interviews. The collection of this information poses a potential threat to the confidentiality of individual data. These risks will be mitigated by several methods, first the study questionnaires will contain little information that would generally be considered to be sensitive, secondly data collection will be conducted in a private place, and finally there is no stigma associated with malaria infection status in the study communities. Malaria is a widely recognized disease in Sierra Leone and seeking treatment is encouraged.

There is also minimal risk associated with the ProCCM intervention. HTR CHWs will continue to provide routine iCCM services. The only change is that in arm 1, HTR CHWs will be paid an extra incentive to proactively visit households twice a month instead of only once every two months and efforts will be made to prevent stock outs in arm 1 and arm 2.

This study will be of direct benefit to study participants in arm 1 and 2 of the study. All participants in the intervention arms will be proactively screened for malaria, and in study arm 1 and 2 the study team will work to ensure a continuous supply of RDTs and ACTs. As an indirect benefit, the study findings may also inform future improvements in the availability of community-level malaria services.

### 5.2 Informed consent

Prior to the start of the study, community meetings will be conducted in each of the communities selected for participation in the study. Community consent will be sought from community leaders for participation in the study. CHWs will follow their standard procedure for seeking verbal consent from households prior to conducting a passive or proactive household visit.

Written, informed consent will be sought from participants in the focus groups, in-depth interviews, and cross-sectional surveys. All information regarding the participants will remain confidential to the extent allowed by law. Unique numerical identifiers will be used for data entry and will not be linked to any personal identifiers other than household GPS coordinates.

### 5.3. Ethical approvals

The study protocol will be submitted to Sierra Leone Ethics and Scientific Review Committee for review and approval prior to the start of study activities. Research approval will also be sought from PATH, Tulane University, and CDC. Research permission will also be sought from respective ministries, regional and district authorities.

## 6. Data management and analysis

### 6.1. Data management

Before the start of data collection, the data collection tools will be piloted and adjustments will be made to the tools. The cross-sectional survey will be programmed in Research Electronic Data Capture (REDCap) on mobile phones or tablets. The design of the forms on the electronic data collection tools will limit the entry of certain data types to enhance quality. For example, dropdown forms will be created in the electronic tools to limit selection to only the information needed.

For data storage and security, encryption technologies will be used and the data will be stored in secure, password-protected servers. Access to this data will be restricted to authorized personnel only and regular backups will be maintained. An initial review will be conducted as part of the data cleaning process to identify inconsistencies and missing values, followed by outlier analysis, and checks for completeness.

After initial data collection, data will be anonymized by replacing identifiers such as name with a unique code. The exception is the enumeration dataset which will be anonymized once the final cross-sectional survey is complete. All data will be archived in a secure repository for a minimum period of five years. The Principal Investigator will have the overall responsibility for data management; Data Manager will oversee data storage, cleaning, and initial analyses; and Field Supervisors will manage quality control and assurance during data collection. Routine Health Information System Data (RHIS) will also be collected for each PHU to identify and compare the numbers of CHW visits across study arms as well as drug stock out information for each CHW/PHU. This data will not contain any human subject or personal identifiable data fields.

Focus group discussions and in-depth interviews will be closed to outside observers and will be held in a private location. Transcripts of all discussions and interviews will be created in Microsoft Word and all personally identifying information will be removed from the study transcripts. Interviews will be translated to English language during the process of transcription. The recordings will be archived with limited access to only the research team; the recordings will be destroyed after the data analysis has been completed. Data coding and analysis will be completed using a qualitative analysis software.

### 6.2. Quantitative analysis

A detailed statistical analysis plan will be produced for the trial. The primary analysis for estimating the treatment effect between intervention and control arms for the primary outcome (proportion of children promptly seeking care for a febrile illness with a qualified provider) will be based on intention-to-treat at the individual level without adjustment for anticipated confounding variables as these are considered to be balanced due to randomization. For all analyses, standard errors of effect estimates will be estimated with a random intercept at the cluster level to account for correlated observations at the cluster level as a result of the community randomized control trial study design. While the primary outcome will be the difference in care-seeking at the final cross-sectional survey, we also plan an interim analysis of the survey data conducted at the cross-sectional survey post ProCCM initial implementation. This measure will be used to understand the temporal dynamics of the effect of the intervention on the study endpoint but is not part of the assessment of the primary outcome which is measured later in time. This analysis would also focus on the primary outcome but these analyses are not considered to be a component of the sustainable increase in care-seeking behavior required to achieve the primary outcome of the trial, and as such no adjustment to the primary outcome analysis is made to account for this extra analysis. Significance testing for these analyses will follow the same principles as the analysis of the primary outcome data.

The primary outcome, prevalence of prompt care-seeking at qualified providers (on the same day or next day after fever onset) among participants aged 6-59months will be analyzed using a multi-level (variance components model) constructed on a generalized linear model framework with a Bernoulli likelihood and a logit link function. Random intercepts will be included for each study cluster and study arm will be included as a fixed effect.

### 6.3. Qualitative analysis

Transcripts from the interviews and focus groups with community members will be reviewed using thematic analysis. An initial set of themes will be defined based on the available literature and the theory of change for each of the different interview groups (community members, HTR CHWs, health facility staff, and DHMT). Additional, emergent themes will be identified and defined through the review of the transcripts. Each theme will be assigned a code and relevant portions of text will subsequently be assigned codes by those conducting the analysis. The coded data will be organized according to study arms. When coding is complete, a synthesis of data within each code will be drafted. Patterns among themes and across types of respondents will be identified and interpreted.

### 6.4. Cost-effectiveness analysis

Data from cost will be analyzed by calculating the incremental cost of delivering the intervention components in each study cluster. Capital items (those with useful lifetimes greater than one-year) will be discounted over their useful life using a 3% discount rate in base case analysis. Additional cases of prompt care-seeking will be estimated using the primary ITT analysis outcome by utilizing the fitted regression model to estimate the counterfactual scenario (*e.g.* no intervention) in intervention clusters.

These estimates will be scaled by population size to account for the sampling fraction in the study area, and further scaled by the estimated rate of febrile illness in the study area to account for the cross-sectional nature of the outcome. The resulting estimates of additional febrile cases seeking treatment will be paired with cluster level costs in order to estimate the incremental cost effectiveness ratio per additional febrile case seeking prompt care from a qualified provider. Uncertainty in these estimates will be addressed in several manners. Primarily these will include one way and scenario analysis in order to incorporate parameter uncertainty in ICER estimates. Additionally, PSA will be used to incorporate joint parameter uncertainty and sampling variation. Finally, ICER estimates will be calculated using a two-stage bootstrapping procedure which involves replicate bootstrap samples of cost and outcomes at both the cluster and individual level, with ICER estimates being recalculated for each bootstrap sample. The resulting estimates will be summarized using cost-effectiveness acceptability curves.

## 7. Timeline

**Figure 6:**
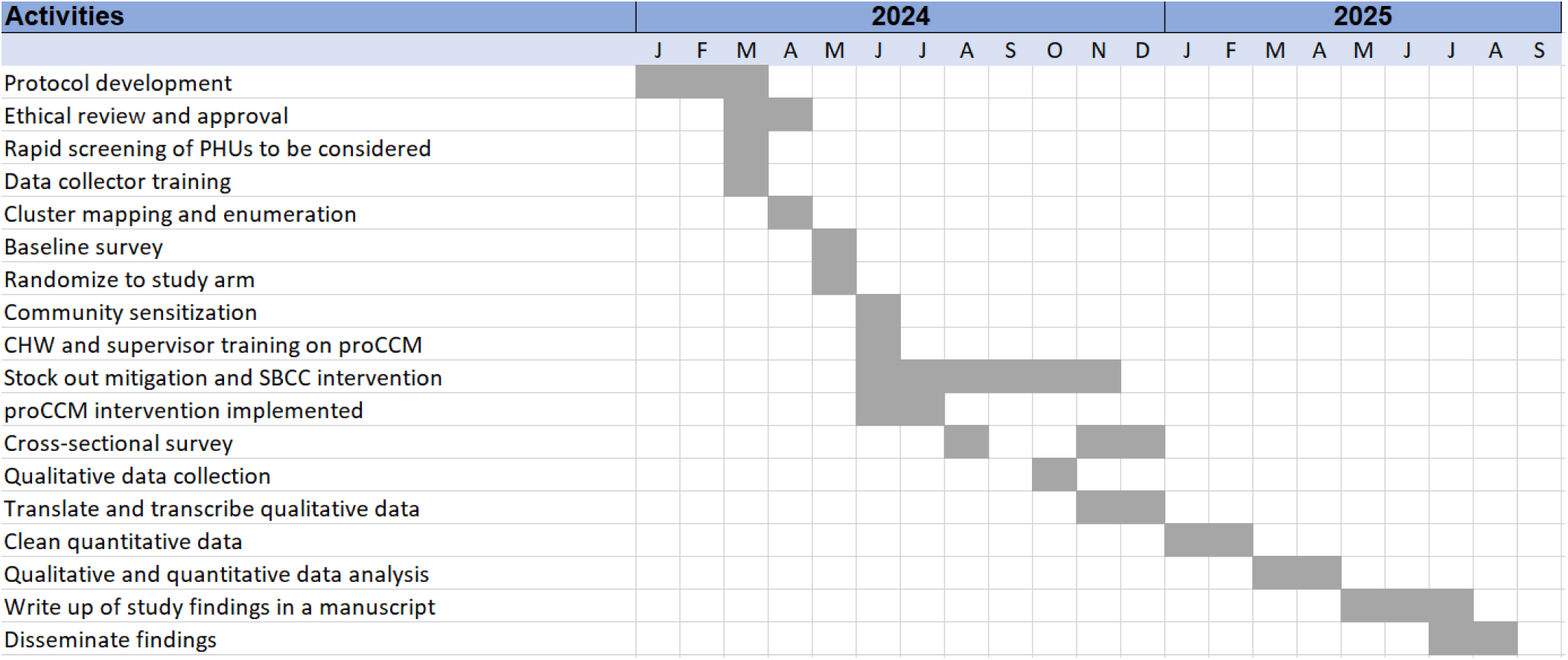
Study timeline

## Appendices

Provide separate attachments.

Each attachment should have a footer/header with the document name, version number and date.

## A short period of proactive community case management (ProCCM) to improve early care-seeking for fever in Sierra Leone: Statistical Analysis Plan

### Section 1: Administrative Information

### Title and trial registration

A short period of proactive community case management (ProCCM) to improve early care-seeking for fever in Sierra Leone

### Trial registration numbers

Clinicaltrials.gov registration number: NCT06395207

Registration date: First Posted (2024-05-02)

### Statistical Analysis Plan version

SAP version number 0.0 September 14, 2024

### Protocol version

Protocol Version 1.0 September 2024

### SAP revisions

| Version Number | Revisions on Previous | Date of revision | Timing of Revision in relation to study | Rationale |
| --- | --- | --- | --- | --- |
| Version 0.0 | NA | September 28<sup>th</sup> 2024 | Prior to midline survey completion | First Version |

### Roles and responsibilities

| Name | Affiliation | Role in SAP |
| --- | --- | --- |
| Joshua Yukich | Associate Professor, Tulane University School of Public Health and Tropical Medicine | Lead SAP author |
| Annie Arnzen | PATH | SAP Contributor / Lead Study Coordinator |

### Signatures

#### Person writing the SAP

Joshua Yukich

#### Senior statistician responsible

Joshua Yukich

#### Chief investigators/clinical leads

Sulaiman Lakoh signed:

Ginika Egesimba signed:

#### Co-investigators

Josh Yukich Annie Arnzen Thom Eisele Temitayo Labor

Abdul Mac Falama Shelby Cash Anne Linn

Annette Ofodum Bailah Molleh Mohamed Jalloh

### Section 2: Introduction

#### Background and rationale

Malaria case management that includes testing and treatment of people experiencing signs and symptoms of illness is an essential strategy in the fight against malaria in many countries in Africa. The utilization and impact of these services is largely dependent on people seeking care when they or their children have fever. In many malaria-affected countries, trends in care­ seeking have not seen the same marked improvements that have been observed in other intervention coverage indicators^1^. The barriers to care-seeking are well documented including the cost of health services, travel time and distance, and negative perceptions regarding the quality of care available^2^•^3^.4

In recent years, many countries have introduced and scaled up community case management (CCM) of malaria (and, in many settings, integrated community case management of childhood illness or iCCM, which includes malaria, pneumonia, and diarrhea) through community health workers (CHWs). The availability of malaria services at the community level through CHWs addresses geographic barriers to care and increases access to life-saving services^5^. However, the utilization of CHWs varies widely by context^1^•^6^. To date, CHW programs have not consistently addressed the known barriers to care such as user fees for services, inadequate CHW supervision, provision of care only from fixed sites, and stock outs of essential medicines and test kits^7^•^8^. Drivers of community utilization of CHWs include trust, satisfaction, and perception of CHW skills and knowledge (Breakthrough ACTION presentation on CHW Literature Review, *unpublished).* The review also found that repeated, proactive visits by CHWs to houses has a positive impact on community utilization of CHW services.

To reduce the gap in care-seeking and address some of the known barriers to seeking care and utilizing CHW services, proactive case finding strategies have been implemented in some countries to increase the prompt treatment of uncomplicated malaria and prevent such cases from developing into severe disease. One such strategy is proactive community case management (ProCCM). In this intervention, CHWs visit households in their communities to proactively offer a defined package of services, such as CCM for malaria or iCCM. In models of this intervention that include a malaria component, CHWs screen for persons with fever and offer malaria diagnostic and other assessment services, as well as malaria treatment, health communication, and referral services.

ProCCM has been shown to be well accepted by the community, feasible to implement, and increases the number of patients treated by CHWs^9,10^. In some settings, the implementation of ProCCM has been linked to improvements in early care-seeking for fever^9^. In Senegal, the number of malaria cases detected and treated by CHWs increased during periods between ProCCM rounds in one observational study^10^. he largest increase in care-seeking during passive periods was seen during the first year of implementation, and continued, albeit at lower levels, during year two and three of the intervention scale-up^10^. A study in Mali also found notable improvements in prompt care-seeking for febrile illness in the context of a ProCCM program with the rate of early effective antimalarial treatment of children under five increasing from 14.7 percent to 35.3 percent^11^. However, a systematic review conducted by Whidden et al found that while many studies of proactive case-finding home visits by CHWs increased access to care and reduced child mortality, there was no significant impact on care-seeking behavior^7^. The certainty of the evidence available to date on the relationship between ProCCM and care-seeking is low and requires further research with rigorous study design.

#### Context in Sierra Leone

Malaria remains a significant public health problem in Sierra Leone and is the leading cause of illness and death among children under five years of age. According to the 2021 Malaria Indicator Survey, malaria prevalence is 22 percent among children aged 6-59 months^12^. Malaria transmission in Sierra Leone is perennial with seasonal peaks occurring during the early rainy season in May and again in October at the end of the rainy season. The primary malaria prevention efforts include universal coverage with insecticide treated nets (ITNs), intermittent preventive treatment in pregnancy (IPTp), and perennial malaria chemoprevention (PMC).

Health services are delivered through a three-tiered system including peripheral health units (PHUs), district/secondary hospitals, and national/regional referral hospitals^13^. Sierra Leone has a large private health sector.

The community health program serves as an extension of the PHUs. In 2022, after a two-year hiatus, the CHW program introduced an integrated approach to scale up malaria diagnosis and include an integrated package of services at the community level^14^. CHWs in easy-to-reach (ETR) areas serve a catchment population of 500-1,000 in 100 to 170 households located between three to five kilometers from the nearest PHU. ETR CHWs are responsible for providing health counseling and making referrals to the PHU. Hard-to-reach (HTR) CHWs serve a catchment population of 300 to 350 in 50 to 60 households in areas that are more than five kilometers from the nearest PHU or between 3km and 5km from the nearest PHU in areas with difficult terrain^14^. CHWs in ETR and HTR areas are expected to proactively visit households in their community once every two months to provide SBCC messages, and preventive and promotional services. However, SBCC messages are not consistently delivered in all areas and CHWs are not trained on all key messages included in the national message guide for malaria. In addition to health prevention and promotional services, HTR CHWs are trained and supplied to provide expanded iCCM services, including diagnosis and treatment of malaria, diarrhea, and pneumonia. However, HTR CHW stock outs are a known barrier to providing reliable testing and treatment services.

The Sierra Leone 2021 Malaria Indicator Survey found that advice or treatment was sought for 75 percent of children who had fever within the preceding two weeks, however only 40 percent sought care on the same or next day following fever onset^12^. Despite the availability of CHWs, only 7 percent of caregivers who sought care for a child with recent fever reported seeking care from a CHW, with the majority seeking care from a government health facility (68 percent)^13^.

#### Rationale for the study

Despite the critical importance of timely identification and treatment of malaria, many families do not promptly seek care for febrile illness for a variety of reasons. In Sierra Leone, ProCCM household visits may serve to improve the visibility of CHWs to the community, increase the community’s level of awareness of CHW capabilities, strengthen the community’s trust in CHWs and, therefore, increase the community’s utilization of CHW even during periods when ProCCM household visits are not occurring.

### Primary and secondary objectives

The primary objective of this study is to:

The secondary objectives are to:

#### Study endpoints

| Outcome | Data collection method | Definition |
| --- | --- | --- |
| The proportion of children 0-59 months with recent fever (within the past two weeks) who sought care on the same day or day after fever onset<br>(note that the formal definition of the study outcome is given in section 6) | Cross-sectional survey | • The numerator will be defined as those children currently 0-59 months who sought care for fever on the same day or next day after symptom onset at a provider identified as a CHW or a public health facility.<br>• The denominator will be all children currently 0-59 months who stayed in sampled households the night before the survey whose caregiver reports have a current fever or had a history of fever in the previous 14 days. |
| Cost per additional fever case promptly seeking care at qualified provider | Cross-sectional survey, project records, DHIS2/CHIS, and provider interviews | • Cost of delivering ProCCM, incremental SBCC and stockout management interventions from the provider perspective will be the numerator<br>• The denominator of the ratio will be the incremental number of fever cases receiving prompt management by a qualified provider. |
| Description of care-seeking behavior for febrile illness of people of all ages and how ProCCM impacts this behavior. | Focus groups | • Barriers and enablers of care-seeking for febrile illness among people of all ages and how ProCCM impacts this behavior as reported by community members |
| Description of how ProCCM impacts community members' perceptions of the availability, cost, quality, and ease of seeking care for febrile illness. | Focus groups (arm 1 only) and in-depth interviews | • Perception of community members and the role of ProCCM in care-seeking described using themes arising from the qualitative study |
| Description of how ProCCM impacts CHW job satisfaction and overall workload | Focus groups | • Job satisfaction and perceived workload as described by CHWs during focus groups and interviews |
| Description of how ProCCM impacts health facility staff, peer supervisor, and DHMT linkages and touchpoints with CHWs. | In-depth interviews | • Differences in touchpoints and connection between CHWs and health facility staff as described through qualitative analysis. |

### Section 3: Study Methods

#### Trial design

1. Optimized standard of care for CCM plus ProCCM - ProCCM will be implemented for two months near the start of the high transmission season, and the existing program will be supplemented to mitigate stock outs of malaria commodities among HTR CHWs and ensure HTR CHWs are adequately trained on SBCC messages to ensure activities are implemented as designed.
2. Optimized standard of care - as in arm 1 without ProCCM
3. Routine implementation (control)- no changes (business as usual) to iCCM, SBCC and stock management.

### Randomization

#### Randomization Plan

Randomization will be conducted by the trial statistician following the baseline survey. Randomization will be conducted using a restricted randomization approach whereby each of the clusters will be randomly allocated to one of the three study arms (1:1:1) by selection of an acceptable randomization from a large number of proposed randomizations, each of which meets a pre-determined set of restriction criteria. Restriction criteria for each cluster for balancing the study arms will be based on data obtained from the baseline survey and from the local malaria control program. Actual criteria used are listed below. The main restriction criteria will be balance in prompt care-seeking between arms. The results of the randomization will be shared with the public during community sensitization meetings.

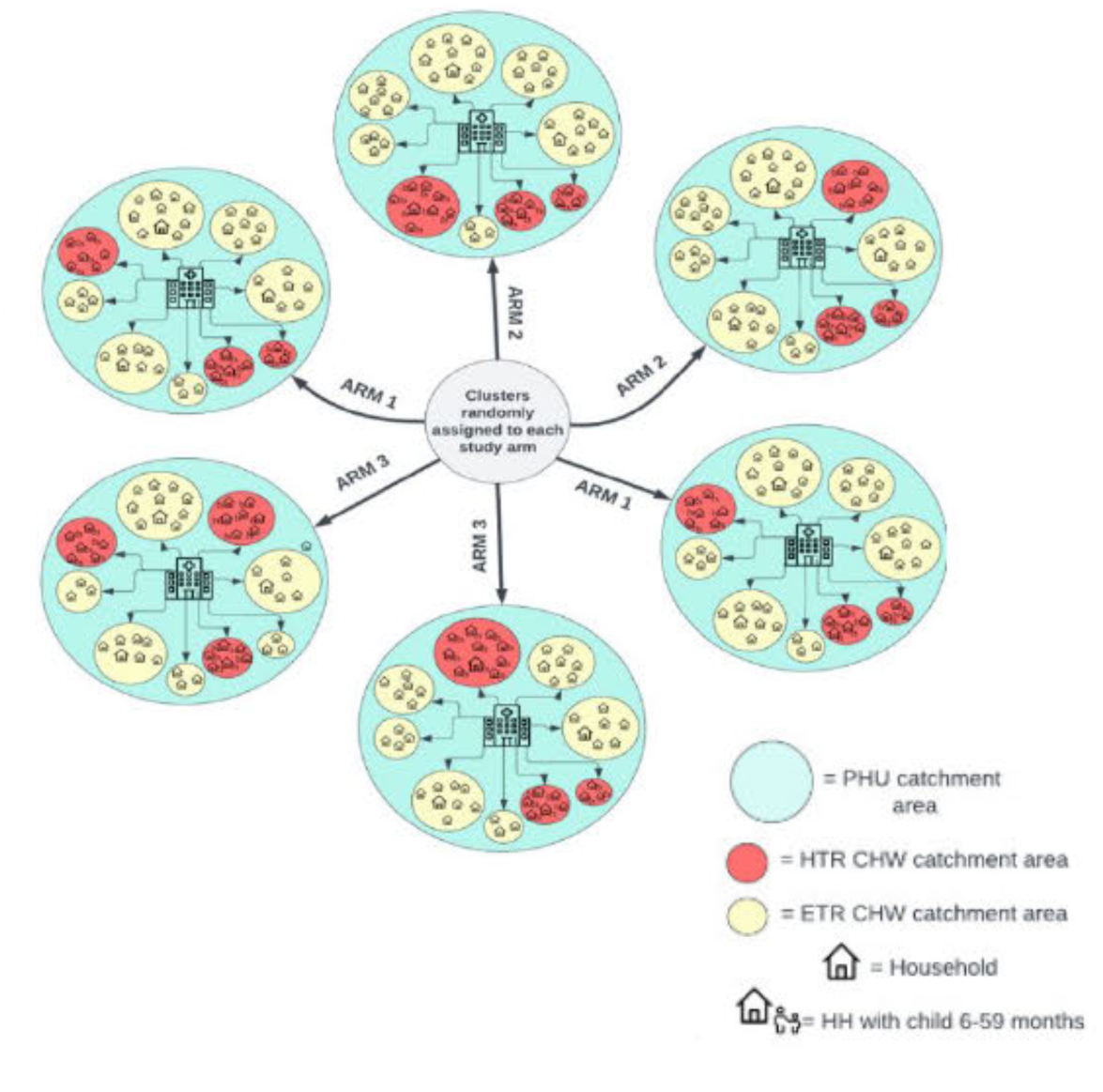

#### Randomization Results

Baseline data was used alongside supplemental information about the study clusters in order to conduct a restricted randomization procedure to allocate study clusters to one of the three study arms in a 1:1:1 ratio. The baseline dataset was used to calculate cluster level summaries of the following variables: proportion of eligible children with a recent fever who sought care on the same or next day after fever onset, and the mean distance from the household location to their PHU location. The enumeration dataset was used to calculate the number of households located in HTR communities in the PHU, and finally an additional supplemental dataset acquired from Population Services International (PSI) was used to identify the number of trained CHWs working in HTR areas in the study clusters and to identify any study clusters without a PSI validated trained CHW.

The restricted randomization procedure was implemented as follows: After creation of a cluster level dataset containing summary measures of each of the above indicators, 70,000 proposed allocation sequences of 45 clusters to three arms were generated, these allocation sequences were then checked against the following criteria:

1. There could be no more than 5 percentage points in difference between prompt care seeking between the arms
2. Differences in the proportion of care seeking could not be considered statistically significant between any pair of arms using a t-test on the cluster summary estimates with p-value < 0.05.
3. The standard deviation of the cluster level care seeking estimates cannot be different by more than 0.07 between arms.
4. The mean of the cluster level estimates for the average distance of a person in the baseline survey from their PHU cannot differ between arms by more than 0.5km
5. The mean of the population size of each cluster (in terms of HHs) cannot differ by more than 50 between arms.
6. The total number of CHW on the validated PSI list cannot differ by more than six CHWs between arms
7. The number of communities with no PSI listed CHW cannot differ by more than one community between arms.

Checking the 70,000 allocation sequences against these criteria resulted in 1,106 sequences which met all criteria. These sequences were then tested for validity, meaning that each cluster must be independently assigned to a study arm, and as such no two clusters should always appear in the same arm or always in separate arms. Additionally, validity requires that the frequencies of any pair of clusters appearing together should be similar across all pairs of clusters. The 1,106 sequences generated in this study met these criteria for validity. Once validity of the list of allocation sequences was established, we selected one allocation at random from the list. The assignment of the three groups to specific study arms was done by taking all six possible permutations for assignment of three arms to three groups (e.g. A= 1, B = 2, C= 3 vs A= 2, B = 1,C= 3) to the integers one through six. The head of the NMCP in Sierra Leone then rolled a fair six-sided die to choose the specific allocation. This resulted in assignment of optimized standard of care plus ProCCM, optimized standard of care and routine implementation to the specific groups established by the random allocation sequence. The results of the randomization yielded the following study arm level estimates at baseline.

Table 3 indicates that the randomization procedure balanced the study arms on the pre-determined study cluster characteristics.

**Table 3:** results of the randomization.

| Study Arm | Prompt Care-Seeking | Mean Distance from PHU (km) | Mean # of HH per cluster | # of HTR CHWs | # of Clusters with no HTR CHW |
| --- | --- | --- | --- | --- | --- |
| Optimized standard of care plus ProCCM | 54.0% | 4.2 | 182 | 40 | 2 |
| Optimized standard of care | 55.9% | 4.0 | 174 | 38 | 1 |
| Routine implementation | 55.0% | 3.8 | 149 | 41 | 1 |

In addition to assessing balance between arms, the baseline survey was used to preliminarily assess assumptions that were used in the sample size calculation. Briefly, sample size was calculated following the approach outlined as follows: a bespoke MonteCarlo simulation algorithm which simulated a three-armed controlled trial with a binary outcome assuming a logistic random effects data generating model was used. We assumed a coefficient of variation k ∼ 0.3 and used the Bonferroni correction for multiple testing given the trials three arms require two tests to determine differences between arms. This results in a p-value of <0.025 being required in a two-sided test to control type-I error at the 5% level. We estimated that the study would have >80% power to detect an increase in early care-seeking from 40% to 60%, with at least 10 children reporting fever in the past two weeks identified in each cluster (assuming 15 clusters per arm, or a total of 45 clusters).

In the baseline survey, the calculated cluster level coefficient of variation (k) on the primary outcome was estimated to be 0.25, or slightly lower than that assumed in the original survey. If the result holds true at later surveys for outcome assessment, the power of the study will be slightly improved. Our sample size calculations also assumed that we would have at least 10 children per cluster reporting fever. The mean number of children reporting fever in each cluster in the baseline survey was approximately 24, substantially higher than anticipated. While the period prevalence of fever is likely to vary over seasons and time, this too should result in improved power if similar fever prevalence is found in outcome assessment surveys. Finally, while the original sample size calculations were based on proportion of prompt care seeking of 40% in the control arm, the baseline survey found 55%, substantially higher than was presumed in sample size calculations. This will result in reduced power to detect a 20 percentage point change in prompt care seeking since a 20 percentage point change from 40% to 60% is a larger relative change than a 20 percentage point change from 55% to 75%. On balance, the baseline survey indicates that the study is adequately powered to detect slightly smaller change than 20 percentage points. With a baseline prompt care-seeking level of 55%, the study is expected to have greater than 80% power to detect a change of 17 percentage point increase, from 55% to 72%, with a p-value < 0.025 assuming k = 0.25 and a sample of 24 children with fever per cluster. The assumptions used for sample size calculations would be considered either very accurate or pessimistic in all cases. The results do not indicate a need for a change in approach for surveys after the baseline.

#### Cross-sectional household survey

A random sample of 60 (70 or the full number of eligible HHs in the midline survey) households in each cluster with children between 6 and 59 months of age will be drawn from the enumeration data set.

Baseline data collection will employ a structured survey instrument consisting of a screening questionnaire to determine household eligibility (*e.g.* presence of child between 6-59 months of age with a history of fever in the preceding 14 days), a consent procedure, and a full caretaker interview for households in which a child with a history of fever is present. The questionnaire will be essentially identical to that used for the outcome assessment surveys and will be administered to caretakes of children with a history of fever or a knowledgeable adult representative for the child and caretaker. The instrument for the full interviews will capture information about the child, the household, as well as other demographic and socio-economic information including perceptions of care seeking and typical practices for febrile children. The screening visit is expected to take no more than 20 minutes, while the full interview could last up to one hour.

#### Survey eligibility

- Household is located within at HTR CHW catchment area of a selected study cluster
- Adult respondent consents to participate in the baseline survey
- A child between 0 and 59 months of age is a *de jure* resident of the household and has a history of fever within the previous 14 days.

#### Procedures for Cross Sectional Surveys

- Households expected to have a *de jure* resident between the ages of 0 and 59 months will be sampled from all HH enumerated in each study cluster using simple random sampling.
- Sampled households will be visited by the survey team. Information about the survey will be provided to the head of household or his/her representative and a brief screening will be administered. If the household head and/or his/her representative is not available at the time of the first visit, the household will be visited up to two additional times at varying times of day to limit non-response. Sampled households that cannot be contacted within three visits will not be included in the study, and they will not be replaced with another household.
- If the sampled household is considered eligible after the screening questionnaire (*i.e.* there is a child in the household between the ages of 0 and 59 months who has a history of fever within the previous fourteen days and a caretaker or representative capable of answering questions for the HH), then a full consent and interview will be conducted in consenting households.
- A questionnaire will be administered to the caretaker of the child or his/her representative and will include questions about household demographics, housing characteristics, malaria prevention, and fever treatment-seeking behavior. The questionnaire will use electronic data capture administered using tablets.

#### Framework

The trial follows a superiority framework. The comparisons will consist of two-sided tests of the null hypotheses that the outcomes in Arm 1 or Arm 2 are statistically indistinguishable from the outcomes in the control arm (Arm 3). All primary comparisons will consist of comparisons of the outcome in the intervention arm vs. the outcome in the control arm.

### Statistical interim analyses and guidance

#### Stopping for harm

The trials do not include formal stopping rules based on harm the risk to trial participants is expected to be minimal; thus formal harm-based stopping rules are not needed. However, this does not preclude investigators or ethics committees stopping the trial for harm should unforeseen consequences of the intervention or trial procedures lead to harms.

#### Interim Analysis

No interim analysis is planned in the study, since the first endpoint is being tested after the first midline survey and statistical testing will be done after each round. Pooled endpoint analysis may be conducted after the endline survey is complete.

#### Timing of final analysis

The final analyses will be conducted first for each survey round separately immediately following each survey, and pooled and sub-group analysis will be conducted collectively after completion of the endline survey.

### Timing of outcome assessments

#### Primary and secondary efficacy outcomes

##### Primary outcome

The primary outcome measure is the prevalence of prompt care-seeking at qualified providers (on the same day or next day after fever onset) among participants aged 0-59 months (The definition is specified in full in a later section). This will be assessed among children aged 0 months than 59 months whose caregiver or survey respondents report a history of fever in the previous 14 days. These outcomes will be ascertained during cross sectional survey visits to randomly selected eligible households.

##### Secondary outcomes

Secondary outcomes are either cost-effectiveness or qualitative and are not covered in this SAP.

### Section 4: Statistical Principles

#### Confidence intervals and *p*-values

The trial is generally intended to control type-I error to less than 5%. Given the multiple tests of primary outcome planned due the three-armed nature of the study type-I error will be controlled using a Bonferroni correction as discussed in more detail under multiplicity section. The main trial results (intervention efficacy estimates) will be presented with 95% confidence intervals and *p*-values.

#### Adherence and protocol deviations

Since the intervention is deployed on a group basis rather than individually, adherence definitions take account of this. Standard adherence will be defined as intention to treat a cluster of residence with the assigned intervention. Cluster level adherence measures will be defined and pre-categorized prior to final analysis and used to categorize the per-protocol trial population.

Standard adherence will be defined as intention to treat a cluster of residence with the assigned intervention. The per-protocol analysis populations will be defined as those living in intervention clusters where the appropriate intervention was deployed according to the planned schedule. Clusters where less than 50% of anticipated intervention visits occurred or where substantial deployment of interventions into control areas occurred (*e.g*. ProCCM visits conducted in control areas) will be removed from the per-protocol analysis population.

Standard protocol deviations will be considered reportable/summarizable when clusters refuse ProCCM despite having been assigned to arm 1 and providing initial study consent. Additionally, protocol deviations will be considered to have occurred. If ProCCM visits by the study team are delayed by more than four weeks from the expected timeline according to study planning. Protocol deviations related to failure to carry out other study procedures such as outcome assessment on a standardized schedule will not be considered reportable unless they affect an entire cluster and result in a delay of primary outcome assessment of greater than one month.

Protocol deviations related to failure to deploy interventions will be summarized in the final trial reports as well as incorporated into the calculation of adherence.

#### Analysis populations

There are two analysis populations for the primary outcome assessment: These are the intention to treat population and the per-protocol analysis population. The intention to treat population consists of all eligible individuals recruited and consented to participate in the study. The primary analysis will be conducted on the intention to treat population. Per-protocol analysis populations will be those eligible, recruited and consented individuals whose adherence cluster level meets the adherence standard.

#### Multiplicity

Whilst the trial tests multiple secondary outcomes, no adjustment will be made for multiplicity due to secondary outcomes because the trial has a single primary outcome. However, as the trial has three arms the Bonferroni correction for multiple testing will be used. Given that the trial’s three arms require two tests to determine differences between the two intervention arms and the control arm, this results in a *p*-value of <0.025 being required in a two-sided test to control type-I error at the 5% level across both tests.

### Section 5: Trial Population

The target population for the intervention consists of all *de facto* and *de jure* residents present in intervention and control clusters during the study period and the CHWs who serve these communities. The population to be sampled for outcome assessment considers several additional inclusion and exclusion criteria as outlined below. First the procedure for cluster establishment is described. Clusters will be established at all HTR CHW locations within a PHU. Prior to baseline data collection, at least 45 clusters will be established across study arms by identifying PHUs and mapping HTR CHWs around PHUs. Cluster boundaries will be established through participatory mapping with HTR CHW and all households within the catchment areas of the HTR CHW in chosen or potential clusters will be enumerated and mapped. Attempts were made to ensure that clusters are geographically separated to minimize the risk of contamination due to CHWs serving households outside of their established service areas. PHU that present significant logistical or operational challenges for data collection, and/or had very small populations of HTR areas, will not be included in the final set of study clusters, or in one case were pooled with a nearby PHU for creation of a cluster of sufficient population size for inclusion.

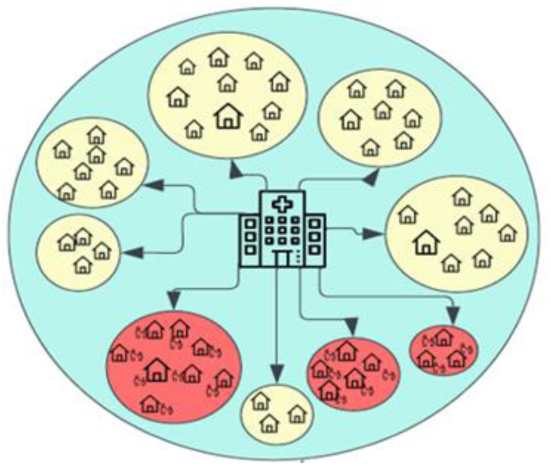

#### Screening data

Each study cluster was enumerated (only households in the catchments of HTR CHWs) prior to a baseline survey and randomization for all HTR CHW areas located within the cluster (PHU). The enumeration consisted of confirming whether any children under five lived in a household, documenting the number of children under five years of age living in each household, and taking GPS locations for all households located within the HTR areas of the PHU (cluster).

#### Enumeration eligibility

Enumerated baseline and randomization established a list of potentially eligible households for outcome assessment (e.g. households who had a *de facto* or *de jure* resident under five years of age).

#### Eligibility

- Eligibility for participation is described in detail in the protocol but in short, household is located within a HTR CHW catchment area of a selected study cluster
- Adult respondent consents to participate in the baseline survey
- A child between 0 and 59months of age is a *de jure* resident of the household and has a history of fever within the previous 14 days.

#### Procedures for Baseline Cross Sectional Survey

- Households expected to have a *de jure* resident between the ages of 0 and 59months will be sampled from all HH enumerated in each study cluster using simple random sampling (stratified by study cluster).
- Sampled households will be visited by the survey team, screened for the presence of an age-eligible child with a history of fever in the previous 14 days.
- If the sampled household is considered eligible after the screening questionnaire (*i.e.* there is a child in the household between the ages of 0 and 59 months who has a history of fever within the previous fourteen days and a caretaker or representative capable of answering questions for the HH), then a full consent and interview will be conducted in consenting households.

#### Recruitment

Recruitment into the study is conducted by first completing an enumeration of all households and their members in the study clusters. This enumeration will be used as a sampling frame to select households with eligible individuals for the cross-sectional surveys. Within each study cluster, a simple random sample of households with potentially eligible children will be selected, eligible children with a history of fever in the previous 14 days as established during screening will be asked to participate in the cross-sectional survey after providing informed consent. Further details of recruitment are contained in the trial protocol.

The CONSORT diagram will include at minimum the following elements.

**Table 4:** CONSORT diagram contents.

| **Cross sectional study (each round)** |
| --- |
| Number of study clusters (by arm) |
| Number of sampled houses (by arm) |
| Number of eligible houses (by arm) |
| Number of consenting houses (by arm) |
| Number of completed interviews (by arm) |
| Number of children with fever history (by arm) |
| Number of incomplete HH surveys (by arm) |
| Number of identified eligible children who did not have the primary outcome assessed (by arm) |

#### Withdrawal/follow-up

It is anticipated that there will be approximately 5-10% attrition per survey round (due to children previously age-eligible aging out of the cohort). This is accounted for in sample size calculations. Level of non-participation in the household surveys is expected to be less than 20%. Non-participation and attrition due to aging out/movement will be summarized by arm and by cluster.

#### Baseline patient characteristics

The study anticipates summarizing several baseline participant characteristics at the individual, household and cluster level. The following table lists these minimum baseline participant characteristics and the expected summary measures which will be reported by arm.

**Table 5:** Baseline participant characteristics (Table one contents)

| Characteristic | Cross-sectional summary measure |
| --- | --- |
| **Cluster Level** |  |
| **Number of clusters** | N |
| **Cluster Size** | Mean N HH (TOTAL HH) |
| **Cluster Size** | Mean N residents (Total N) |
| **Baseline Treatment Seeking** | Proportion of children aged 0 – 59 months with a history of fever in previous 14 days who sought care on the same or next day after fever onset. |
| **Household Level** |  |
| **HH size** | Mean N residents (SD) |
| **Bednet ownership** | Proportion HH with $\geq 1$ Mosquito Bednet |
| **Bednet ownership** | Proportion HH with $\geq 1$ Mosquito Bednet per 2 residents |
| **Individual Characteristics** |  |
| **Age** | Mean (SD) |
| **Sex** | Proportion female |
| **HH size** | Mean hh size of included hh (SD) |
| **Bed Net Use** | Proportion of children under 5 years of age with a fever in the past 14 days who slept under a night the night before the survey |

### Section 6: Analysis

#### Outcome definitions

The primary outcome measure is the proportion of children aged 0-59 months who have a history of fever within the previous 14 days who sought treatment or advice from any provider on the day of or the day after symptom onset. Fever will be defined as any current or reported history of fever reported by the caregiver/taker or representative to have occurred within the previous 14 days. For those children who are both age eligible and have a positive report of fever history, questions will be asked to the caregiver/taker as to whether or not care/treatment or advice was sought for the fever. If care/treatment or advice was sought, the timing of the first care/treatment or advice seeking attempt in relation to the onset of the febrile symptoms will be ascertained. Prompt care seeking will then be defined as seeking care on the same day or the day after symptom onset, excluding where care was solely sought from a church, mosque, drug shop/market stall, drug peddler, traditional healer or any combination of these non-formal providers.

#### Analysis methods

##### Primary outcome

The primary unadjusted analysis will be conducted on the intention to treat analysis population without adjustment for any anticipated confounding variables as these are considered to be balanced due to randomization. The analysis of the primary outcome, proportion of children under five years of age with a fever in the previous 14 days who sought care on the day of or the next day from symptom onset, will be analyzed using a multi-level (variance compartments model) constructed on a generalized linear model framework with a bernoulli likelihood and a logit link function. Random intercepts will be included for each study cluster and study arm will be included as a fixed effect coded categorically as 0 for arm 3 and 1 for arm 2 and a second indicator variable coded as 0 for arm 3 and 1 for arm 1. The model will take the form below where *pij* is probability of positivity at the individual level (*i* indexes individuals within clusters and *j* indexes clusters), *α* is the global intercept, *X*ij is the arm assignment for individual *i* in cluster *j*, *βarm* is the arm effect to be estimated, *uj* are random intercepts for the cluster and *σ* is the standard deviation of the random intercept distribution:

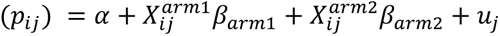

Where the likelihood is of the form:

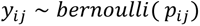

And the random intercepts are assumed to follow a normal distribution:

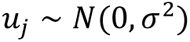

Model results will be presented as the estimates of *α* and the odds ratio above and the standard deviation or variance of the random effects distribution. 95% confidence intervals for the odds ratio and *α* estimates as well as *z*-statistics and *p*-values for each coefficient will be presented.

#### Covariate adjusted analysis of the primary outcomes

Adjusted analyses will be carried out on the analysis of the primary outcomes to determine whether the estimate of treatment-effect is affected with the inclusion of additional covariables. The prespecified covariates will be developed and tested prior to final analysis. One additional analysis will include all covariables which are used in restricted randomization with measures treated exactly as specified in randomization. These variables are shown below in the table 6, along with other covariables which may be considered in adjusted analysis. These analyses will be prespecified for the primary outcome prior to data lock and the statistical analysis plan will be updated to reflect these analyses. Examples of pre-specified covariates that may be included in the adjusted analyses are described in table 6 which will be finalized prior to data lock.

**Table 6:** Proposed Covariables.

| Variable | Categorization (if applicable) | Analysis Population | Restricted Randomization Variable |
| --- | --- | --- | --- |
| Baseline prompt care seeking | Calculated at cluster level | ITT, per-protocol | Yes |
| Mean Distance from assigned Primary Health Unit | Calculated at cluster level | ITT, per-protocol | Yes |
| Number of Households in cluster | Cluster Level | ITT, per-protocol | Yes |
| Number of CHW in HTR areas in Cluster | Cluster Level | ITT, per-protocol | Yes |
| Month | Calendar Months Post Intervention or by Survey Round | ITT, per-protocol | No |
| Age | Age in Months | ITT, per-protocol | No |

#### Subgroup analysis of the primary outcome

We will perform a series of subgroup analyses according to the list subgroups in the table below. Imputation for these baseline missing covariates (see section *Missing Data*) will be carried out before categorizing. Assessment of the homogeneity of treatment effect by a subgroup variable will be conducted by inclusion of the treatment, subgroup variable, and their interaction term as predictors in the adjusted models of primary outcome, and the *p*-value presented for the interaction term. If the *p*-value is less than 0.05, we will present separate effect estimates and confidence intervals for each category of the sub-group variable.

**Table 7:** Planned sub-group analyses.

| Subgroup Name | Categorization | Rationale |
| --- | --- | --- |
| Gender of Head of Household | Male vs. Female | Behavioral and occupational differences may act as effect modifiers; to demonstrate equity of the intervention effect |
| Gender of Respondent | Male vs. Female | Gender of child may act as effect modifiers to the intervention effect on care seeking; to demonstrate equity of the intervention effect |
| Survey Round | Primary analysis is of each survey round data independently. | The duration and timing of any effect from ProCCM may depend on the time since intervention. |
| Age | <= 11 months of age vs 12 - 59 months of age | Behavioral differences by age may act as effect modifier |
| Baseline Treatment Seeking | High vs. low (>= median cluster treatment seeking vs. < median cluster treatment seeking) | Areas with high baseline levels of treatment seeking may respond to the ProCCM intervention differently than |
|  |  | those with lower levels of baseline treatment seeking |
| Mothers' Education | Some Formal Education vs. No Formal Education | Mothers or caregivers with more formal education may respond differently to the ProCCM intervention than those without formal education |
| Proximity to PHU | =< Median distance from PHU within a cluster vs. > median distance to PHU within a cluster | Lower barriers to care seeking in terms of travel time and cost may lead to different responses to ProCCM |

### Additional Analyses

#### Pooled Analysis using control and supply chain support only arms as control

Should no statistically significant difference between the control arm and the supply chain support only arm be found in primary analysis, these two arms will be pooled for an analysis of the effect of ProCCM compared to the pooled outcomes of all individuals in both of these arms. This analysis will follow similar statistical principles to the primary analysis specified above, with the exception that the indicator variable for the supply chain only arm will be removed from the model.

### Missing data

#### Missing outcome data

Significant effort will be made to reduce missing outcome data by revisiting sampled households multiple times and pre-scheduling follow up visits where possible. When missing data does arise due to failed outcome assessment no imputation will be used. Missing outcomes due to participant absence will result in removal of the observation from the model.

#### Missing co-variates

Missing covariates (as defined in the SAP prior to data lock) will be imputed using simple imputation methods in the covariate adjusted analysis based on the covariate distributions, should the proportion of missing values for a particular covariate be less than 5%. For a continuous variable, missing values will be imputed from random values from a normal distribution with mean and standard deviation calculated from the available sample. For a categorical variable, missing values will be imputed from random values from a uniform distribution with probabilities *P*1, *P*2, … *P*k from the sample. Seed for the imputation will be pre-set as an 8-digit number based on the date of analysis and documented in all scripts relying on pseudo-random number generators.

#### Harms

The main risks associated with the intervention are the risk of disclosure of confidential information or loss of privacy during interviews. As these harms are not related to the intervention no formal plan for statistical analyses of harms to study participants. Unexpected harms may occur during the course of trial and will be considered for reporting to ethics boards though no formal analysis is planned.

##### Statistical software

Reporting of statistical analysis will include specific details of software platform, including language, version and details of any additional libraries used in analysis. It is anticipated the most analyses, data cleaning and preparation will be conducted using the R language.

## Data Availability

All data produced in the present study are available upon reasonable request to the authors

